# Using network analysis to illuminate the intergenerational transmission of adversity in the ALSPAC cohort

**DOI:** 10.1101/2021.12.11.21267654

**Authors:** Chad Lance Hemady, Lydia Gabriela Speyer, Janell Kwok, Franziska Meinck, G.J. Melendez-Torres, Deborah Fry, Bonnie Auyeung, Aja Louise Murray

**Author notes:** contributed equally to this paper (Bonnie Auyeung, Aja Louise Murray). Corresponding author: Chad Lance Hemady, School of Social and Political Sciences, University of Edinburgh, United Kingdom. Email addresses of authors: CLH LGS JK FM GMT DF BA ALM.

## Abstract

**Objective:** The effects of maternal exposure to adverse childhood experiences (ACEs) may be transmitted to subsequent generations through various biopsychosocial mechanisms. However, studies tend to focus on exploring one or two focal pathways with less attention paid to links between different pathways. Using a network approach, this paper explores a range of core prenatal risk factors that may link maternal ACEs to infant preterm birth (PTB) and low birthweight (LBW).

**Methods:** We used data from the Avon Longitudinal Study of Parents and Children (ALSPAC) (n = 8 379) to estimate two mixed graphical network models: Model 1 was constructed using adverse infant outcomes, biopsychosocial and environmental risk factors, forms of ACEs, and sociodemographic factors. In Model 2, ACEs were combined to represent a threshold ACEs score (≥ 4). Network indices were estimated to determine the shortest pathway from ACEs to infant outcomes, and to identify the risk factors that are most vital in bridging these variables.

**Results:** In both models, childhood and prenatal risk factors were highly interrelated. Childhood physical abuse, but not threshold ACEs, was directly linked to LBW. Further, exposure to second-hand smoke, developing gestational hypertension, prenatal smoking, first time pregnancy, not being White, and older age were directly linked to LBW, while developing gestational diabetes, having previous pregnanc(ies), and lower educational attainment were associated with PTB. Only pre-eclampsia was directly linked to both outcomes. Network indices and shortest pathways plots indicate that sexual abuse played a central role in bridging ACEs to other risks and poor infant outcomes. Overall, prenatal smoking was determined as the most influential bridge node.

**Conclusions:** As child physical abuse was directly linked to low birthweight, and child sexual abuse and prenatal smoking were the most influential bridge nodes, they can be considered priority candidate targets for interventions to disrupt intergenerational risk transmission. Further, our study demonstrates the promise of network analysis as an approach for illuminating the intergenerational transmission of adversity in its full complexity.

## Background

One of the United Nations’ sustainable development goals (target indicator 3.2) for 2030 is to reduce preventable deaths of neonates and children under 5 years of age globally (1). Preterm birth (PTB) (births before 37 weeks of gestation) and low birthweight (LBW) (birthweight <2500 grams) remain among the leading causes of child morbidity and mortality (2,3) and are associated with both short- and long-term consequences throughout the lifespan (4). The pathogenesis of these adverse birth outcomes has been linked to a number of interconnected and potentially interacting prenatal risk factors, such as maternal exposure to teratogenic agents (e.g., tobacco, illicit substances, air pollutants), psychological distress, psychosocial stressors (e.g., financial difficulties), and biological complications (e.g., pre-eclampsia, infections) (4–7). Prenatal life is a critical phase for fetal programming and a highly sensitive period for both maternal and fetal biology (8,9). Thus, exposure to the aforementioned risk factors may cause alterations in the gestational environment which could result in deficits in fetal development (4,7–9).

Maternal exposure to adverse childhood experiences (ACEs), characterized by forms of maltreatment (e.g., physical abuse, neglect) and household dysfunction (e.g., parental incarceration), have also been directly linked to both infant outcomes; however, the results are inconsistent (10–13). The prenatal risk factors mentioned above, along with genetic, epigenetic and other environmental factors, are posited as pathways of transmission of ACE-related sequelae from mother to infant (8,14). For example, ACE exposure may lead to the biological embedding of toxic stress and in turn impair the immune-inflammatory system (15). Increased systemic inflammation may lead to increased risk of intrauterine infections, which is one of the most important mechanisms leading to PTB and LBW (4,7,16). However, studies tend to focus on exploring one or two focal pathways with less attention paid to the interconnectivity and co-occurrence of these individual-, interpersonal-, and macro-level risks. Drawing from graph theory and network science, network analysis is a statistical technique in which conditional independence relationships of all variables are modelled (17,18). Through this, the interrelationships between exposure variables can be estimated which may lead to the identification of unique interrelations among variables that cannot be identified in traditional analyses. Such relations are, for example, more challenging to discern in techniques such as multiple regression analyses, which tend to examine the relation between one focal outcome and multiple predictor variables while largely ignoring the shared relations between predictors (18). Compared to regression models, network analysis is a more integrative approach through which complex multidimensional chains of risks can be analyzed.

The aim of this study is therefore to explore pathways through which intergenerational transmission of adversity may operate by estimating network models that allow for insights into the complex interrelationships among many variables. Network models are commonly used in psychology to abstract connections among symptoms (19). They are also used in epidemiologic studies to identify risk factors for disease outcomes (20–23) and predict the spread of infectious diseases (24).

Despite its promise, thus far, only a few studies have used a network approach to investigate the interrelatedness of ACEs (25) and the relationships between ACEs and poor health outcomes in adulthood (25–29). For instance, Breuer and colleagues used network analysis to investigate the direct relations between ACEs and adult mental ill-health. They found high connectivity between all ACEs, and identified neglect and domestic violence exposure as influential factors that mediate connections between ACEs (27). They also reported that childhood sexual abuse was the only link between ACEs and adult mental ill-health. These findings suggested that neglect and domestic violence exposure are optimal candidate targets for interventions against ACEs while interventions against sexual abuse are more likely to prevent negative long-term mental ill health. Similarly, network analysis was used to unravel complex relationships between ACEs and differential effects of distinct ACEs on various personality dimensions (26). These authors found that childhood physical abuse and maternal rejection played a central role in shaping personality, specifically, the former was negatively associated with cooperativeness and positively associated with novelty seeking, while the latter was negatively associated with novelty seeking and self-directedness and positively associated with harm avoidance. Similarly, Pereira-Morales and colleagues examined the relations between multiple risk factors for mental illness in adulthood (22). They found a relation between sexual abuse in childhood and higher levels of perceived distress in adulthood, and a direct link between emotional neglect and problematic alcohol consumption in adulthood. Overall, worry, perceived distress, and low energy were identified as the risk factors that played a central role in affecting mental health. In contrast with previous studies that used a selective approach (single ACE) or a cumulative approach (using the de facto ≥ 4 ACEs threshold score) to examine relations between ACEs and outcomes, network models have demonstrated a much sparser association pattern and at the same time illuminated complex relations between multiple risks and outcomes (22,27). To our knowledge, no study has used a network approach to examine intergenerational transmission of adversity using multiple biopsychosocial pathways.

## Methods

### Study design and population

This study was a secondary data analysis of the Avon Longitudinal Study of Parents and Children (ALSPAC) dataset. Pregnant women resident in Avon, UK with expected dates of delivery 1st April 1991 to 31st December 1992 were invited to take part in the study. The initial number of pregnancies enrolled is 14,541 (for these at least one questionnaire has been returned or a “Children in Focus” clinic had been attended by 19/07/99). Of these initial pregnancies, there was a total of 14,676 foetuses, resulting in 14,062 live births and 13,988 children who were alive at 1 year of age(30,31). After excluding participants without obstetric records, with multiple pregnancies (twin, triplet, etc.), and still births, the final sample was 8 379. Project details, data documentation, and specific details on ethical approval are available on the ALSPAC study website: https://www.bristol.ac.uk/alspac. Further, please note that the study website contains details of all the data that is available through a fully searchable data dictionary and variable search tool: http://www.bristol.ac.uk/alspac/researchers/our-data/.

### Variables and Measurement

#### Outcomes

Infant preterm birth (PTB) and low birthweight (LBW) were derived from obstetric records and recoded into dichotomous variables (yes/no).

#### ACEs

Maternal ACEs were measured using a 31-item checklist and 12-item checklist as part of self-reported questionnaires gathered at 32 weeks gestation and 33 months postpartum, respectively (32,33). Ten items were used to represent types of ACEs: physical, emotional, and sexual abuse, physical neglect, emotional neglect, parental incarceration, parental death, parental separation, parental divorce, and having had a parent with mental illness. The ALSPAC Study Team (33) recoded these items into dichotomous variables (yes/no). For this analysis, parental death, separation, or divorce were aggregated into one dichotomous variable (yes = if ≥ 1 domain was experienced), which follows the World Health Organization guidelines for coding ACEs (34). Finally, a threshold (≥ 4) ACEs variable was derived by computing the ACE sum score and collapsing it into a dichotomous variable (yes/no). This was done for comparison with previous research that commonly takes this approach to defining ACE exposure (35).

#### Prenatal risk factors

Fifteen variables, gathered between 18- and 32-weeks gestation through self-completion questionnaires, were used to represent potential prenatal risk factors.

#### Psychosocial risk factors

Two items were used to represent psychosocial risk factors: exposure to discrimination and financial difficulties. Mothers were asked if they think they had been unfairly treated in the past year due to their sex, skin color, family background, religion, the way they dress, the way they speak, or other reasons, respectively. Discrimination score was recoded into a dichotomous variable (yes/no) (33). To represent financial difficulties, mothers were asked to rate the difficulty of affording food, clothing, heating, rent, and things they need for the baby, respectively, using a 4-point scale (‘very difficult’ to ‘not difficult’). Sum scores were derived with values ranging from 0 = ‘no financial difficulties’ to 15 = ‘maximum financial difficulties’. Sum score values were coded as missing if any of the 10 component items were missing (33).

#### Psychological risk factors

The 10-item Edinburgh Postnatal Depression Scale (EPDS) was used to evaluate depressive symptoms in the past week. This measure has been validated and has demonstrated high sensitivity and specificity (36) and the Cronbach’s alpha for the current sample exceeded 0.80 (37). The response categories for each item ranged from 0 = ‘quite often’ to 3 = ‘never’. Sum scores were derived with values ranging from 0 = ‘not depressed’ to 29 = ‘very depressed’. Sum score values were coded as missing if any of the 10 component items were missing (33). Prenatal anxiety was measured using the 8-item anxiety subscale (feeling panicky, upset for no obvious reason, strung-up inside, like going to pieces, as though they might faint, uneasy and restless, worrisome, and having upsetting bad dreams) of the validated Crown Crisp Experiential Index (CCEI). Item-level response categories ranged from 0 = ‘never’ to 4 = ‘very often’. A composite measure (ranged from 0 = ‘not anxious’ to 16 = ‘very anxious’) was derived with values coded as missing if at least one of the component items was missing.

#### Environmental risk factors

Three items were used to represent environmental risk factors. At 18 weeks gestation, mothers were asked if their partner smoked (yes/no) and if any other household member smoked (yes/no). Additionally, one item measured passive smoke exposure at 32 weeks gestation (‘how often during the day [are you] in a room or enclosed place where other people are smoking?’) using a 6-point scale with response categories ranging from: ‘all the time’, ‘>5 hrs’, ‘3-5 hrs’, ‘1-2 hrs’, and ‘not at all’ (33). This variable was recoded into a dichotomous variable (yes/no).

#### Medical risk factors

Five items were used to examine medical risk factors during pregnancy: urinary tract infection (UTI), genital herpes, gestational hypertension, pre-eclampsia, and gestational diabetes. At 32 weeks gestation, as part of the self-completion questionnaires, mothers were asked if they had UTI (yes/no) or genital herpes (yes/no) during the past three months (33). Gestational hypertension was derived from obstetric records and was defined as having an elevated blood pressure (systolic blood pressure >139 mmHg [measured in millimeters of mercury] or diastolic blood pressure > 89 mmHg) on at least two occasions after 20 weeks gestation (38). To note, the median number of antenatal clinic visits were 14 per woman (38). Pre-eclampsia (yes/no) was derived from obstetric records and was defined as being diagnosed with gestational hypertension and concurrent proteinuria (38). Finally, gestational diabetes (yes/no) was derived from obstetric records and was defined as having traces of glycosuria (2+ or more) in the urine on at least two occasions (38).

#### Risky health behaviors

Three items were used to investigate risky health behaviors during pregnancy: prenatal smoking, alcohol, and illicit drug use. At 18 weeks gestation, mothers were asked how often during pregnancy they have taken amphetamine, barbiturate, crack, cocaine, heroin, methadone, ecstasy, or other drugs. This item was recoded into a dichotomous variable (33).

At 32 weeks gestation, mothers were asked how many cigarettes they smoked per day. This item was recoded into four categories (0 = ‘none’, 1 = ‘1-9’, 2 = ‘10-19’, 3 = ‘20+’) (33). Finally, the ALSPAC study team derived the total alcoholic units consumed per week by combining the number of half-pints of beer, glasses of wine, drinks of spirits, and other alcohol drinks consumed per week (ranging from 0 to 65) (33).

#### Covariates

The following *a priori* covariates were included in the analyses: age, educational attainment (ranging from lowest [‘none’] to highest [‘degree’]), ethnicity, and parity (39,40). Highest level of educational attainment ranged from lowest (‘none/CSE’) to highest (‘Degree’). Ethnicity was recoded into a dichotomous variable (1 = ‘White’, 0 = ‘Other ethnicities’), given that 97.3% were White. Finally, parity was recoded into a dichotomous variable (0 = first time pregnancy). See Additional File 1 for the item questions and response categories.

### Analytic strategy

Pairwise Markov Random Field (PMRF) models (41) were estimated to explore the underlying mechanisms of intergenerational transmission of ACEs to PTB and LBW. PMRF models consist of nodes which represent observed variables and edges which represent relations between two nodes after conditioning on all other nodes (41). When represented visually, the thickness of an edge represents the magnitude of the association (strong = thick, weak = thin) while the color (positive = blue, negative = red) represents the sign. The analyses were conducted using R (R Core Team, 2013). Two network models were estimated: the first model (29 nodes) included all variables outlined above, while the second model (22 nodes) included a node which represented the threshold ACEs score instead of individual ACEs. For network estimation, we used the *mgm* (42) method as implemented in the *estimateNetwork* function in the *bootnet* package (43). Mixed graphical models (MGM) were constructed given that the data contained both dichotomous and continuous variables, and were estimated and selected via regularized LASSO (*least absolute shrinkage and selection operator*) nodewise regressions (42,44). The LASSO neighborhood regression limits false positive findings (or spurious edges) by reducing weak edges to zero which leads to a sparse network structure. To further limit spurious edges, a penalty approach was utilized by setting the tuning hyperparameter to *γ* = 0.5, selected using the Extended Bayesian Information Criterion (EBIC), which is typically set between 0 and 0.5 (45). Setting the hyperparameter to 0.5 provides conservative estimates that err on the side of parsimony (18). A drawback of this high specificity is that some edges are potentially not represented in the network.

Edges in MGM models are combined estimates from neighborhood regression (i.e., estimates from regressing node A on B, and vice versa). As such, the *ruleReg* argument of the model estimation function was set to ‘*OR’* to specify that an edge should be included if at least one estimate is non-zero (42). Missing data were once imputed via chained equations using the *mice* package (46). This method provides unbiased parameter estimates on the assumption of missing at random. Node predictability, or how well a node can be predicted by all remaining nodes, were estimated using the *mgm* package (42) and visualized in the network plot. Node predictability is based on variance explained for continuous variables and classification accuracy for categorical variables. The networks were visualized using the *qgraph* package (47) which uses the Fruchterman-Reingold algorithm which places nodes that are more strongly connected closer together.

Centrality indices (strength, betweenness, closeness) are typically used in network analysis to quantify the relative importance of each node in relation to the whole network structure (48). However, given that our aim was to explore the pathways between ACEs and adverse infant outcomes, *bridge* centrality indices were estimated instead. Bridge centralities identify nodes that are most vital in communication between communities (or hubs within the network). The *networktools* package was used to estimate bridge strength, closeness, and betweenness (49). Bridge centrality is estimated by summing the absolute value of all edges that lie between node A and all nodes that are not in the same community as node A. Bridge closeness is defined as the inverse of the average length of the path from node A to all other nodes in a different community as node A. While bridge betweenness quantifies the extent to which node B lies on the shortest path between nodes A and C, where both nodes are from distinct communities (49). Additionally, the shortest pathways that link ACEs to PTB and LBW were visualized using *qgraph* package (47). Finally, to test the accuracy of edge-weights, 1,000 nonparametric bootstraps were performed on the two models, respectively, using the *bootnet* package (43).

## Results

### Descriptive statistics

**Table 1:**
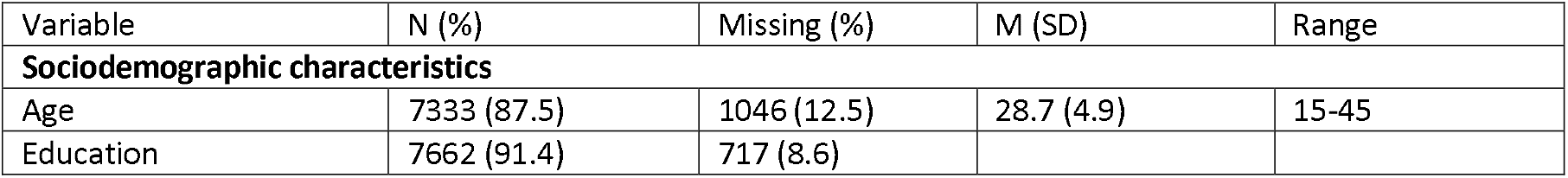
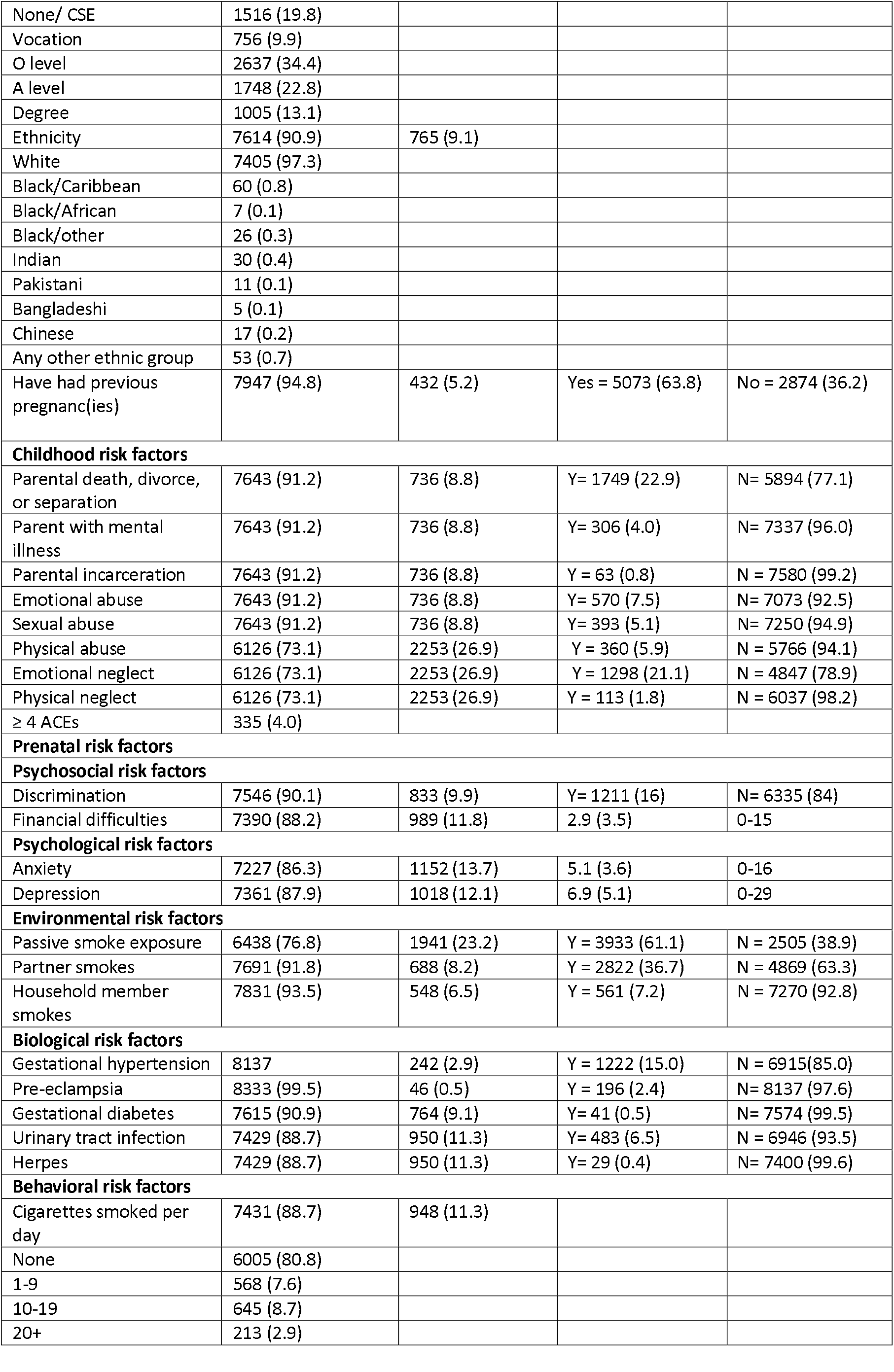
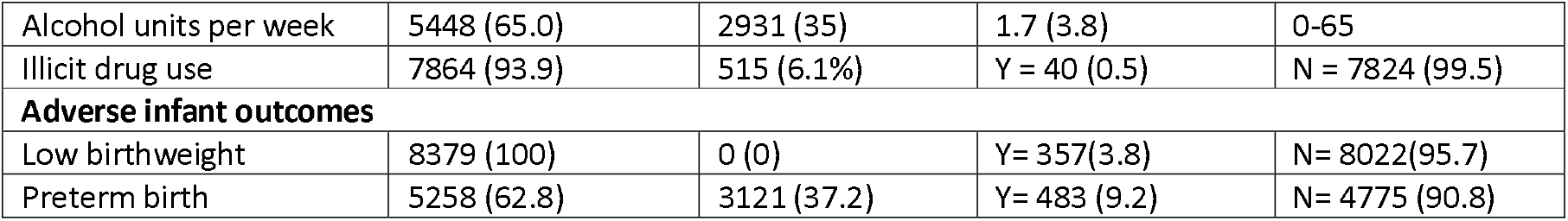
Sociodemographic characteristics of the sample (n = 8 379)

| Variable | N (%) | Missing (%) | M (SD) | Range |
| --- | --- | --- | --- | --- |
| **Sociodemographic characteristics** |  |  |  |  |
| Age | 7333 (87.5) | 1046 (12.5) | 28.7 (4.9) | 15-45 |
| Education | 7662 (91.4) | 717 (8.6) |  |  |
| None/ CSE | 1516 (19.8) |  |  |  |
| Vocation | 756 (9.9) |  |  |  |
| O level | 2637 (34.4) |  |  |  |
| A level | 1748 (22.8) |  |  |  |
| Degree | 1005 (13.1) |  |  |  |
| Ethnicity | 7614 (90.9) | 765 (9.1) |  |  |
| White | 7405 (97.3) |  |  |  |
| Black/Caribbean | 60 (0.8) |  |  |  |
| Black/African | 7 (0.1) |  |  |  |
| Black/other | 26 (0.3) |  |  |  |
| Indian | 30 (0.4) |  |  |  |
| Pakistani | 11 (0.1) |  |  |  |
| Bangladeshi | 5 (0.1) |  |  |  |
| Chinese | 17 (0.2) |  |  |  |
| Any other ethnic group | 53 (0.7) |  |  |  |
| Have had previous pregnanc(ies) | 7947 (94.8) | 432 (5.2) | Yes = 5073 (63.8) | No = 2874 (36.2) |
| **Childhood risk factors** |  |  |  |  |
| Parental death, divorce, or separation | 7643 (91.2) | 736 (8.8) | Y= 1749 (22.9) | N= 5894 (77.1) |
| Parent with mental illness | 7643 (91.2) | 736 (8.8) | Y= 306 (4.0) | N= 7337 (96.0) |
| Parental incarceration | 7643 (91.2) | 736 (8.8) | Y = 63 (0.8) | N = 7580 (99.2) |
| Emotional abuse | 7643 (91.2) | 736 (8.8) | Y= 570 (7.5) | N= 7073 (92.5) |
| Sexual abuse | 7643 (91.2) | 736 (8.8) | Y= 393 (5.1) | N= 7250 (94.9) |
| Physical abuse | 6126 (73.1) | 2253 (26.9) | Y = 360 (5.9) | N = 5766 (94.1) |
| Emotional neglect | 6126 (73.1) | 2253 (26.9) | Y = 1298 (21.1) | N = 4847 (78.9) |
| Physical neglect | 6126 (73.1) | 2253 (26.9) | Y = 113 (1.8) | N = 6037 (98.2) |
| ≥ 4 ACEs | 335 (4.0) |  |  |  |
| **Prenatal risk factors** |  |  |  |  |
| **Psychosocial risk factors** |  |  |  |  |
| Discrimination | 7546 (90.1) | 833 (9.9) | Y= 1211 (16) | N= 6335 (84) |
| Financial difficulties | 7390 (88.2) | 989 (11.8) | 2.9 (3.5) | 0-15 |
| **Psychological risk factors** |  |  |  |  |
| Anxiety | 7227 (86.3) | 1152 (13.7) | 5.1 (3.6) | 0-16 |
| Depression | 7361 (87.9) | 1018 (12.1) | 6.9 (5.1) | 0-29 |
| **Environmental risk factors** |  |  |  |  |
| Passive smoke exposure | 6438 (76.8) | 1941 (23.2) | Y = 3933 (61.1) | N = 2505 (38.9) |
| Partner smokes | 7691 (91.8) | 688 (8.2) | Y = 2822 (36.7) | N = 4869 (63.3) |
| Household member smokes | 7831 (93.5) | 548 (6.5) | Y = 561 (7.2) | N = 7270 (92.8) |
| **Biological risk factors** |  |  |  |  |
| Gestational hypertension | 8137 | 242 (2.9) | Y = 1222 (15.0) | N = 6915(85.0) |
| Pre-eclampsia | 8333 (99.5) | 46 (0.5) | Y = 196 (2.4) | N= 8137 (97.6) |
| Gestational diabetes | 7615 (90.9) | 764 (9.1) | Y= 41 (0.5) | N= 7574 (99.5) |
| Urinary tract infection | 7429 (88.7) | 950 (11.3) | Y= 483 (6.5) | N = 6946 (93.5) |
| Herpes | 7429 (88.7) | 950 (11.3) | Y= 29 (0.4) | N= 7400 (99.6) |
| **Behavioral risk factors** |  |  |  |  |
| Cigarettes smoked per day | 7431 (88.7) | 948 (11.3) |  |  |
| None | 6005 (80.8) |  |  |  |
| 1-9 | 568 (7.6) |  |  |  |
| 10-19 | 645 (8.7) |  |  |  |
| 20+ | 213 (2.9) |  |  |  |
| Alcohol units per week | 5448 (65.0) | 2931 (35) | 1.7 (3.8) | 0-65 |
| Illicit drug use | 7864 (93.9) | 515 (6.1%) | Y = 40 (0.5) | N = 7824 (99.5) |
| **Adverse infant outcomes** |  |  |  |  |
| Low birthweight | 8379 (100) | 0 (0) | Y= 357(3.8) | N= 8022(95.7) |
| Preterm birth | 5258 (62.8) | 3121 (37.2) | Y= 483 (9.2) | N= 4775 (90.8) |

#### Network analysis

For both models, 8 379 participants were included. In general, bootstrapped confidence intervals (CIs) around the edge estimates for the two models were narrow. The bootstrapped accuracy plots are available in Additional file 1. Network visualization are presented in Figures 1 and 2. To facilitate interpretability only edges above 0.2 are visualized. To note, edge weights of MGM models represent regression weights based on standardized data, and the scales of edges differ depending on the type of node. For instance, edges between two continuous nodes represent partial correlations and thus cannot be greater than 1, while edges between binary nodes represent logistic regressions and therefore have no upper limit (44,50). As such, the magnitudes of the binary and continuous edges cannot be directly compared for strength. Finally, node predictability measures were visualized using node rings, with blue rings indicating proportion of explained variance for continuous nodes while purple rings indicate accuracy of intercept model for dichotomous nodes. The red rings indicate additional accuracy achieved by all remaining variables for binary nodes (51).

**Figure 1:**
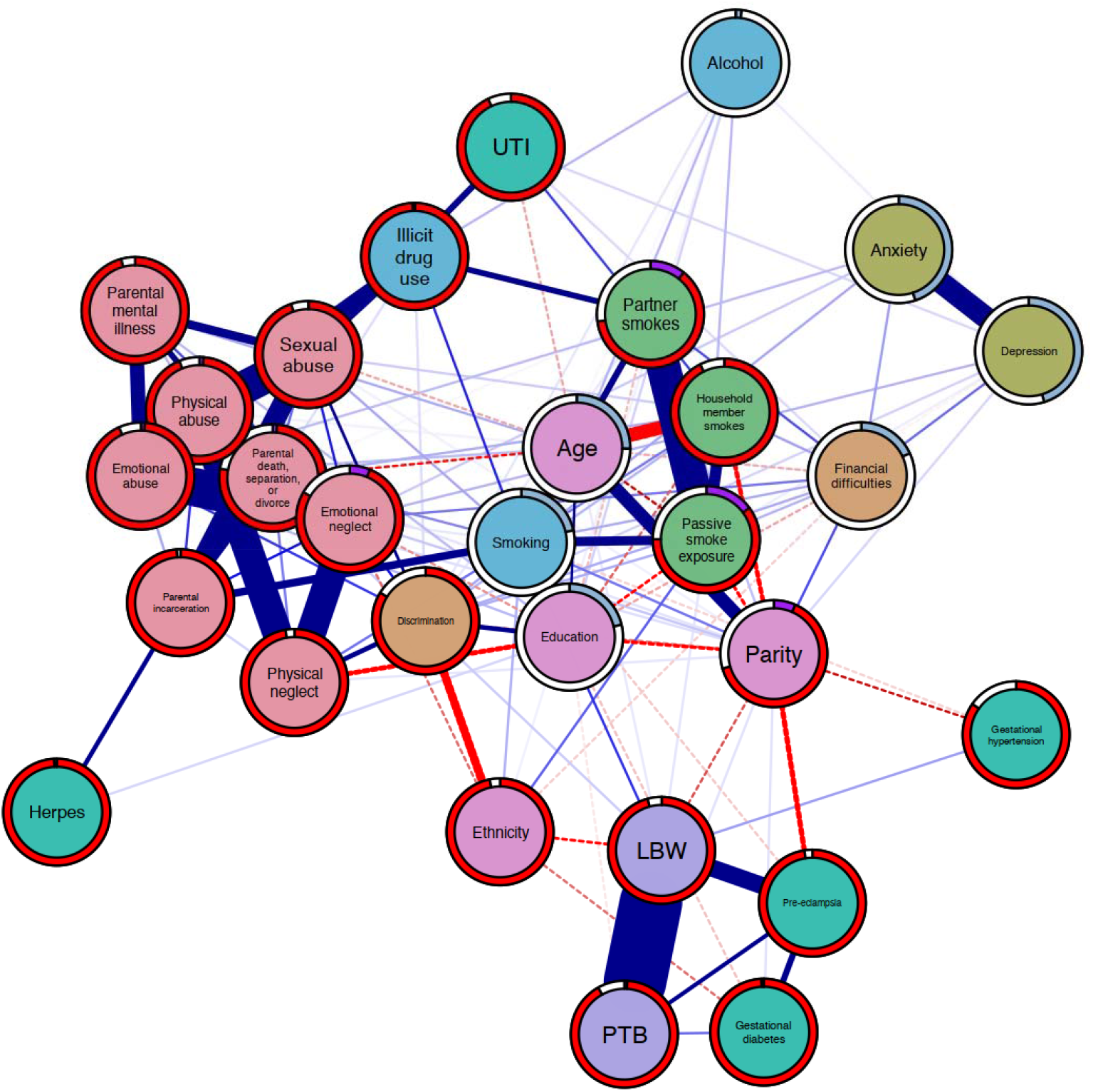
Network displaying the interrelationships between ACEs, a wide array of prenatal risk factors for preterm birth and low birthweight, and the outcomes of interest. Carnation nodes represent ACEs, purple nodes represent the outcomes, teal nodes indicate biological risks, blue nodes indicate risky behaviours, orange nodes indicate psychosocial risks, green nodes represent environmental risks, and pink nodes represent covariates. Blue edges suggest positive association while red edges (dashed) represent negative associations. Node predictability measures visualized using node rings, with blue rings indicating proportion of explained variance for continuous nodes while purple rings indicate accuracy of intercept model for dichotomous nodes. The red rings indicate additional accuracy achieved by all remaining variables.

**Figure 2.**
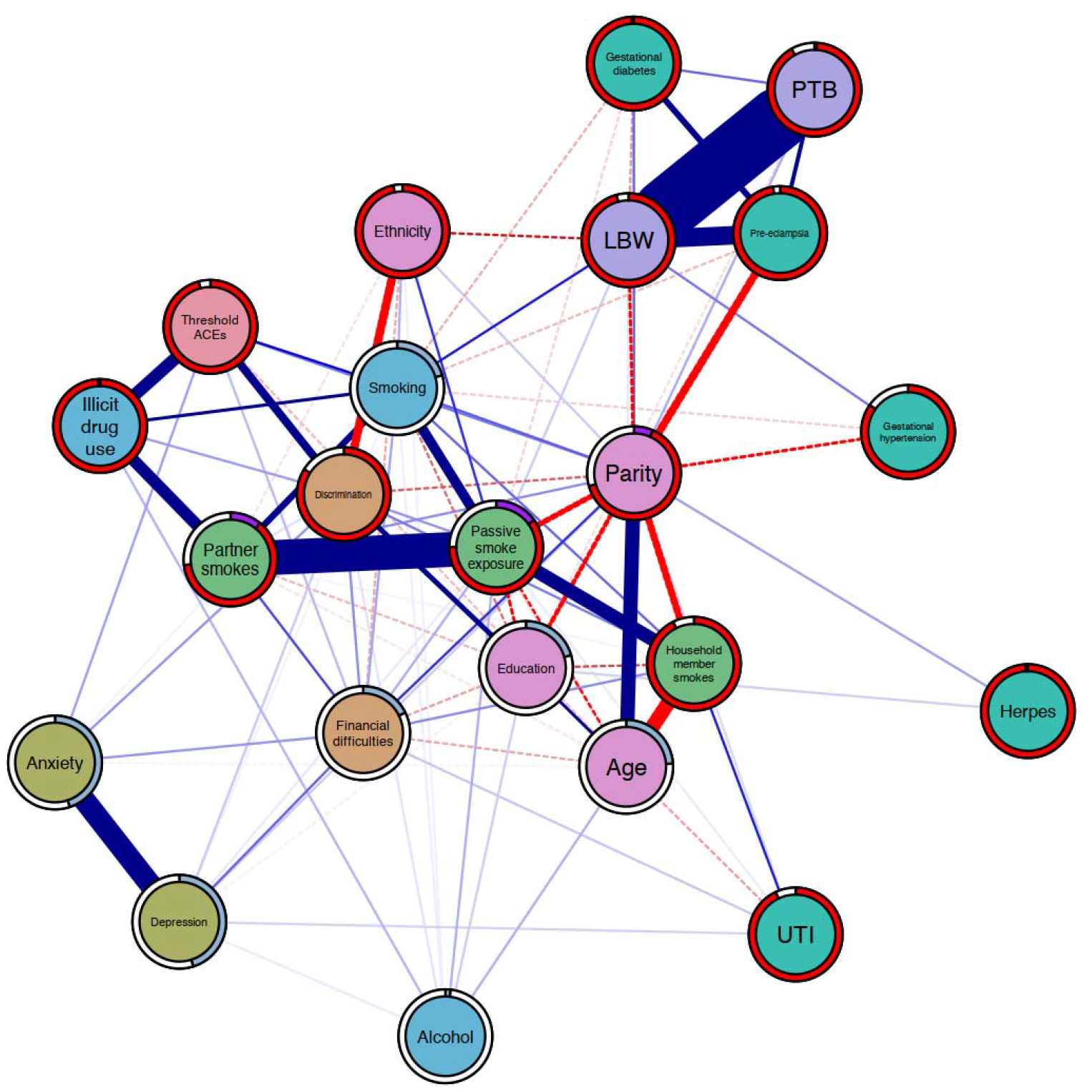
Network displaying the interrelationships between ACEs, a wide array of prenatal risk factors for preterm birth and low birthweight, and the outcomes of interest. Carnation nodes represent ACEs, purple nodes represent the outcomes, teal nodes indicate biological risks, blue nodes indicate risky behaviours, orange nodes indicate psychosocial risks, green nodes represent environmental risks, and pink nodes represent covariates. Blue edges suggest positive association while red edges (dashed) represent negative associations. Node predictability measures visualized using node rings, with blue rings indicating proportion of explained variance for continuous nodes while purple rings indicate accuracy of intercept model for dichotomous nodes. The red rings indicate additional accuracy achieved by all remaining variables.

#### Model 1: individual ACEs

Model 1 estimated high interconnectivity between forms of ACEs, with each one connected to at least five other ACEs. The ACEs characterized by household dysfunction, apart from parental mental illness, were directly linked to at least one medical risk and at least two environmental risks and two risky health behavior nodes. The neglect nodes were directly linked to psychosocial, psychological, and risky health behavior nodes while the abuse nodes were connected directly to at least one psychological, environmental, medical, and risky health behavior nodes, respectively. Older participants were more often exposed to emotional abuse and emotional neglect while younger participants were more often sexually abused.

Overall, there is high interrelatedness among risk nodes with each one connected to at least one other risk. For instance, depression was directly linked to exposure to discrimination and financial difficulties, developing UTI infection, prenatal anxiety, passive exposure to second-hand smoke, and alcohol use. Similarly, both psychosocial risk nodes were connected to all the environmental and psychological risks and at least one risky health behavior node, respectively.

Certain nodes appear to cluster together and create hubs in the network (forms of ACEs, illicit drug use, and discrimination/ environmental risks, smoking, and financial difficulties/ adverse birth outcomes, medical risks, and ethnicity/ psychological risks & financial difficulties). Results from the bridge centrality metrics (Additional file 2: Figure S10) demonstrated that in the ACEs community, sexual abuse has the highest bridge strength, closeness, and betweenness centrality; overall, smoking was determined as the most influential bridge node.

In relation to the infant outcomes, physical abuse was directly linked to LBW but not PTB. In addition, participants who were exposed to passive smoke, developed gestational hypertension, smoked, were not White, had no previous pregnancy, and who were older more often had infants with LBW. While participants who developed gestational diabetes, had previous pregnancies, and had lower educational attainment more often had PTB. Only the pre-eclampsia node was directly linked to both PTB and LBW.

Other forms of ACEs were not directly linked to either outcome but conditionally linked through various pathways. Apart from physical neglect and emotional neglect that both estimated discrimination and ethnicity as the shortest pathway to the LBW node, the general shortest pathway is through the smoking and education nodes (see Additional file 2 for shortest pathway plots: Figure S1-S8).

#### Model 2: threshold ACEs

Model 2 demonstrates an approximately similar pattern with the previous model. The global network structure suggests high interconnectivity among nodes, with all nodes connected to at least two other nodes. Certain communities within the network were also formed (threshold ACEs, illicit drug use, smoking, partner smokes, and discrimination/ adverse birth outcomes & medical risks).

The threshold ACEs node was not directly linked to either infant outcome, but the shortest pathway determined was through the smoking node (Additional file 2: Figure S9). As in the first model, the smoking node was estimated as the most influential bridge node (Additional File 2: Figure S11). Alternatively, developing pre-eclampsia was directly linked to both PTB and LBW. Further, results showed that participants that developed gestational diabetes, had previous pregnanc(ies), and lower educational attainment more often had PTB infants. While participants who were exposed to second-hand smoke, developed gestational hypertension, did not develop gestational diabetes, had no previous pregnanc(ies), were not White, and were older more often had infants with LBW.

## Discussion

Adopting a network approach, we explored the underlying mechanisms that may link maternal adverse childhood experiences to infant preterm birth and low birthweight, while accounting for sociodemographic factors. First, there was high interrelatedness among childhood and prenatal risk factors in both models. This expands on evidence that found that ACEs tend to co-occur (52,53) and are linked to a wide range of persistent biopsychosocial health risks (35,54). Yet, only physical abuse, and not threshold ACEs, demonstrated a direct relationship with LBW. Childhood physical abuse is a prevalent toxic stressor and its far-reaching consequences on health and development is well-established (55). It has been shown that physically abused children exhibit elevated C-reactive protein levels (a biomarker of inflammation), possibly as a way for the body to adapt to physical insults (15). Exposure to prolonged or acute toxic stress during this significant developmental phase may lead to long-term deficits in the immune-inflammatory physiology and may carry over to the gestational biology and cascade to embryonic and fetal development (9).

The bridge metrics indicated that sexual abuse was an influential bridge node (Additional File 2: Figure S10), and the shortest pathway plots (Additional File 2: Figure S1-S8) demonstrated that this node is central in linking other forms of abuse to adverse infant outcomes. The findings are consistent with Breuer and colleagues ACEs network model that showed that sexual abuse played a central role in bridging ACEs and mental disorders in adulthood (27).Childhood sexual abuse is strongly associated with likelihood of exposure to other ACEs and has been identified as the most synergistically reactive form of ACEs, meaning that interactions with other adversities in childhood or adulthood substantially increased health consequences (56). Thus, sexual abuse can be considered an optimal target for intervention because preventing it may increase the likelihood of destabilizing the entire ACE network and may prevent the occurrence of long-term health consequences (27). However, there is limited information on the effectiveness of interventions on preventing sexual abuse. For instance, a recent review by Ligiero and colleagues found that education programs for sexual abuse prevention have been shown to increase knowledge of abuse and promote protective behaviors among school-aged children; yet, additional evidence is needed to assess the impact on the prevalence and incidence of abuse (57). Empowerment and self-defense training for early and late adolescents have shown reductions in sexual violence (57). For instance, a cluster-randomized study in Kenya of a behavior-based sexual assault prevention program, with components of empowerment, gender relations, and self-defense for girls and promotion of healthy gender norms for boys, have shown reductions in sexual assault among girls in this population (58). However, more research on the effectiveness of these types of programs in various contexts is needed (57). It is also important to note that the onus of violence prevention should not be on children, it should be on society and on adults who should protect children. More importantly, prevention programs should target potential perpetrators. Parenting programs to improve parent-child communication have been shown to increase awareness and recognition of sexual abuse and promote positive dialogue, and can play an important role in physical abuse reduction and prevention, but limited evidence is available on actual reductions in sexual violence victimization. (57). Further, despite the numerous international treaties and legislation implemented at the domestic and national level, sexual abuse remain an insidious problem due to lack of political will to enforce these policies.

Second, we found that multiple and individual ACEs were linked to the outcomes through various pathways, and in general, the most common and most influential pathway was prenatal smoking. Exposure to physical abuse and sexual abuse in childhood are risk factors for higher smoking frequency in adolescence and adulthood (59).

Further, there is corroborative evidence that suggest that prenatal smoking is a mediator between ACEs and PTB and LBW, respectively (10,60). Various chains of risk have been posited; for instance, early stressors such as ACEs may negatively impact the development of the neurobiological system (disruption of the dopamine circuit and impairment of stress-regulatory circuits, *inter alia*) (61). To compensate, ACE-exposed children may turn to substances (like nicotine) to stimulate dopaminergic neurons or to modulate stress or negative affect. In turn, early smoking initiation may contribute to drug-seeking behavior, nicotine dependence, and relapse vulnerability that carry over to pregnancy (62,63). To this point, those who have been maltreated as children are at increased risk of developing prenatal depressive or anxiety symptoms (64) and may have greater difficulties in reducing tobacco use during pregnancy (65).

According to the *2017 Global Burden of Diseases, Injuries, and Risk Factors Study*, smoking was among the top risk factors for burdens of disease and mortality among women (66). Relatedly, prenatal smoking has been linked to suboptimal utero-placental oxygen and blood flow and is considered as one of the major risk factors for a wide array of gestational complications, including PTB and LBW (67). Aside from being the most influential risk node, we found that prenatal smoking was directly linked to LBW. This underscores that prenatal smoking is a candidate target for intervention to disrupt intergenerational risk transmission. There is some evidence that pharmacological treatments, coupled with psychosocial interventions, help promote smoking cessation during pregnancy; however, its impact on gestational outcomes are still unknown (68). Behavioral change policies and interventions, which involve addressing negative mental habits or altering the stimuli within the micro-environments to influence health-related behaviors are found to be beneficial (69,70). For instance, evidence suggests that behavioral counseling can support smoking cessation by helping women develop strategies for managing stress or cravings to smoke (70). Given that socioecological factors like having friends or a partner who smoke are barriers for smoking cessation in pregnancy (70,71), addressing norms as cues for behavioral change may be beneficial not only for maternal-fetal health, but the broader public health.

Third, consistent with previous evidence (4,7,72), we found that environmental risks (exposure to second-hand smoke) and sociodemographic factors (ethnicity, age, education) were directly linked to either PTB or LBW. Our results further indicated that women who were not White were more often neglected in childhood, exposed to discrimination, and experienced financial difficulties during pregnancy. Further, women who had lower educational attainment were more often exposed to multiple ACE, had experienced discrimination, financial difficulties, depression, were exposed to second-hand smoke, and used tobacco during pregnancy. As Marmot and colleagues report, even after accounting for socioeconomic disadvantage, minority ethnic groups are more likely than White British people to report poor health (73). Ethnic minority populations, compared to their more advantaged counterparts, are more likely to be subjected to institutional and cultural racism and discrimination; have less access to material resources, education, and health and welfare services, and have poorer living and working conditions (74). These are intersecting and intricately linked social determinants that exacerbate health inequities.

Finally, we found that gestational hypertension was linked to LBW while gestational diabetes was linked to PTB. Pre-eclampsia is the severest hypertensive disorder in pregnancy, and women with pre-existing diabetes are at an increased risk (75). In both models, we found that pre-eclampsia was a risk factor for both PTB and LBW, which is consistent with previous studies (5,72). It is found to decrease utero-placental blood flow and subsequently increase maternal (e.g., cardiovascular and cerebrovascular diseases) and child (e.g., fetal growth restriction, attention deficit/hyperactivity disorder) health complications (76). Antihypertensive pharmacological treatment and routine blood pressure measure at each antenatal appointment is recommended (72,77).

### Limitations

This study is subject to a number of limitations. First, items obtained from self-completion questionnaires are vulnerable to social desirability bias. To address this, validated measures were used where possible. Second, the items related to ACEs were not part of a validated measure nor did they measure other important parameters of risk exposure (i.e., severity, duration, chronicity) which would have provided a more accurate representation. Third, the study did not account for genetic and epigenetic factors which also play a role in birth outcomes.

## Conclusion

By using network analysis, we were able to map out the web of relationships between ACEs and found that when one ACE is present, other types of ACEs are likely to be present as well. Our network also demonstrated the complex interplay between ACEs, prenatal risk factors, and infant preterm birth and low birthweight. Physical abuse was the only childhood adversity linked to low birthweight. This highlights the methodological limitation of using threshold ACE scores and provides evidence that individual ACEs are not equal in the strength of their effect and consequently, undercuts the rationale for using the threshold score in policy and practice settings (56). Finally, childhood sexual abuse and prenatal smoking were identified as the most influential factors in the network and may lend themselves as optimal targets for prevention of intergenerational risk transmission.

## Data Availability

All data produced in the present study are available upon reasonable request to the authors

## List of Abbreviations

ACES: adverse childhood experiences
ALSPAC: Avon Longitudinal Study of Parents and Childs
CCEI: Crown Crisp Experiential Index
CSE: Certificate of Secondary Education
EBIC: Extended Bayesian Information Criterion
EPDS: Edinburgh Postnatal Depression Scale
LBW: low birthweight
LASSO: Least Absolute Shrinkage and Selection Operator
MGM: Mixed graphical models
mmHG: millilitres of mercury
PMRF: pairwise Markov Random Field
PTB: preterm birth
UTI: urinary tract infection

## Declarations

### Ethics approval and consent to participate

Ethical approval for the study was obtained from the ALSPAC Ethics and Law Committee and the Local Research Ethics Committees. Informed consent for the use of data collected via questionnaires and clinics was obtained from participants following the recommendations of the ALSPAC Ethics and Law Committee at the time. The study was carried out in accordance with the Declaration of Helsinki regarding the ethical conduct of medical research involving human subjects.

### Consent for publication

Not Applicable

### Availability of data and materials

<u>The study website</u> contains details of all the data that is available through a fully searchable data dictionary and variable search tool.

### Authors contributions

All authors were engaged in the overall conceptualization, study design, investigation, and findings interpretation. CLH, LGS, and ALM were involved in the analysis. All authors contributed to provide critical review and revision of the manuscript. All authors have reviewed and approved this manuscript.

## Acknowledgements

We are extremely grateful to all the families who took part in this study, the midwives for their help in recruiting them, and the whole ALSPAC team, which includes interviewers, computer and laboratory technicians, clerical workers, research scientists, volunteers, managers, receptionists and nurses.

## Additional file

Additional File 1: Table S1 - Instruments used, questions, and response categories

Additional File 1: Figures S1/S2 - bootstrapped confidence intervals of the edge weights for Model 1 and Model 2

Additional File 1: Figures S3/S4 - Results of the edge weight difference test for Model 1 and 2

Additional File 2: Figures S1-S9 - Shortest pathways plots

Additional File 2: Figures S10/S11 - Bridge centrality indices for Model 1 and 2

