## Additional file 1 for "Using network analysis to illuminate the intergenerational transmission of adversity in the ALSPAC cohort"

Table S1: Instruments used, questions, and response categories

| **Demographic characteristics** | **Question** | **Response categories** | **Timepoint**  **(gestation)** |
| --- | --- | --- | --- |
| Age | Age upon completion of questionnaire |  | 32w |
| Education | What educational qualifications do you have? Please tick all that apply | None/CSE, Vocational, O level, A level, Degree | 32w |
| Ethnicity | How would you describe [your] race or ethnic group | a) White  b) Black Caribbean  c) Black/African  d) Black/Other  e) Indian  g) Pakistani  h) Bangladeshi  i) Chinese  j) Any other ethnic group | 32w |
| Parity | Number of previous pregnancies |  | 18w |
| **Adverse childhood experiences** | **Question** | **Response Categories** | **Timepoint**  **(gestation)** |
| Parental death, separation or divorce | a) Before you were 17 your parent died | Yes, affected me a lot, Moderately affected, Mildly affected, Yes, but did not affect, No | 32w |
|  | b) Before you were 17 your parents separated |  |  |
|  | b) Before you were 17 your parents divorced |  |  |
| Parental mental illness | Before you were 17 a parent was mentally ill | Yes, affected me a lot, Moderately affected, Mildly affected, Yes, but did not affect, No | 32w |
| Parental incarceration | Before you were 17 a parent was imprisoned | Yes, affected me a lot, Moderately affected, Mildly affected, Yes, but did not affect, No | 32w |
| Emotional abuse | Before you were 17 a parent was emotionally cruel | Yes, affected me a lot, Moderately affected, Mildly affected, Yes, but did not affect, No | 32w |
| Sexual abuse | Before you were 17 you were sexually abused | Yes, affected me a lot, Moderately affected, Mildly affected, Yes, but did not affect, No | 32w |
| Physical abuse | Thinking back to your childhood, were you physically abused (e.g., beaten) | Yes, severely; Yes, somewhat; No | child 33 months |
| Emotional neglect | Thinking back to your childhood, did you feel neglected emotionally? | Yes, severely; Yes, somewhat; No | child 33 months |
| Physical neglect | Thinking back to your childhood, were you physically neglected (e.g., not fed or clothed properly) | Yes, severely; Yes, somewhat; No | child 33 months |
| **Prenatal risk factors** | **Question** | **Response Categories** | **Timepoint**  **(gestation)** |
| *Psychosocial risk factors* | |  |  |
| Discrimination score | Do you think you have been unfairly/unjustly treated in the last 12 months because of your sex, your skin colour, the way you dress, your family background, the way you speak, or your religion | Often, Sometimes, Never | 32w |
| Financial difficulties score | How difficult at the moment do you find it to afford these items: food, clothing, heating, rent or mortgage, things you will need for the baby | Often, Sometimes, Never | 32w |
| *Psychological risk factors* | |  |  |
| Anxiety subscale (CCEI) (8 items) | a) Do you feel upset for no obvious reason? | Very often, Often, Not very often, Never | 32w |
|  | b) Do you sometimes feel panicky? |  |  |
|  | c) Do you feel strung up inside |  |  |
|  | d) Do you ever have the feeling you are going to pieces? |  |  |
|  | e) Have you felt as though you might faint? |  |  |
|  | f) Do you feel uneasy and restless? |  |  |
|  | g) Do you worry a lot? |  |  |
|  | h) Do you have bad dreams which upset you when you wake up? |  |  |
|  | b) Do you feel tingling or pricking sensations in your body, arms, or legs? |  |  |
|  | c) Do you feel tired or exhausted? |  |  |
|  | d) Do you feel sick or have indigestion? |  |  |
|  | e) Do you find that you have little or no appetite? |  |  |
|  | f) Do you often have excessive sweating or fluttering of the heart? |  |  |
|  | g) Can you get off to sleep alright? |  |  |
| Depression (EPDS) | Your feelings the past week: | As much as I could, Not quite as much, definitely not so much, not at all | 32w |
|  | a) I have been able to laugh and see the funny side of things |  |  |
|  | b) I have looked forward with enjoyment to things |  |  |
|  | c) I have blamed myself unnecessarily when things went wrong |  |  |
|  | d) I have been anxious or worried for no good reason |  |  |
|  | e) I have felt scared or panicky for no very good reason |  |  |
|  | f) Things have been getting on top of me |  |  |
|  | g) I have been so unhappy that I have had difficulty sleeping |  |  |
|  | h) I have felt sad or miserable |  |  |
|  | i) I have been so unhappy that I have been crying |  |  |
|  | j) The thought of harming myself has occurred to me |  |  |
| *Environmental risk factors* | |  |  |
| Passive smoke exposure | Please indicate how often during the day you are in a room or enclosed place where other people are smoking | All the time, >5 hrs, 3-5 hrs, <1hr, Not at all | 32w |
| Partner smokes | Does your partner smoke? | No, Yes, cigarettes; Yes, cigars; Yes, pipe; Yes, others; Don’t have a partner | 18w |
| Household member smokes | Apart from yourself and your partner, are there any other members of your household who smoke | Yes or No | 18w |
| *Biological risk factors* | |  |  |
| Gestational hypertension | Derived from at least two measures with either a systolic blood pressure > or equal to 140mm or a diastolic pressure > or equal to 90mm | Yes or No | obstetric records |
| Pre-eclampsia | Derived from at least two measures with either a systolic blood pressure > or equal to 140mm or a diastolic pressure > or equal to 90mm plus at least 1+ proteinuria as the raised BP | Yes or No | obstetric records |
| Gestational diabetes | Derived from at least two measures with 2+ or more glycosuria in the urine | Yes or No | obstetric records |
| UTI | Have you had urinary infection in the last 3 months | Yes or No | 32w |
| Herpes | Have you had genital herpes in the last 3 months | Yes or No | 32w |
| Behavioural risk factors | |  |  |
| Alcohol use | Derived from the questions:  In the past week, how many i) half-pints of beer; ii) glasses of wine; iii) drinks of spirits; iv) other alcoholic drinks did you drink? | Frequency count | 32w |
| Smoking | Derived from the question: How many cigarettes do you smoke per day | None, 1-9, 10-19, 20+ | 32w |
| Illicit drugs | Derived from the question: How often have you taken amphetamine, barbiturate, crack, cocaine, heroin, methadone, ecstasy, or other drugs during pregnancy | Yes or No | 18w |

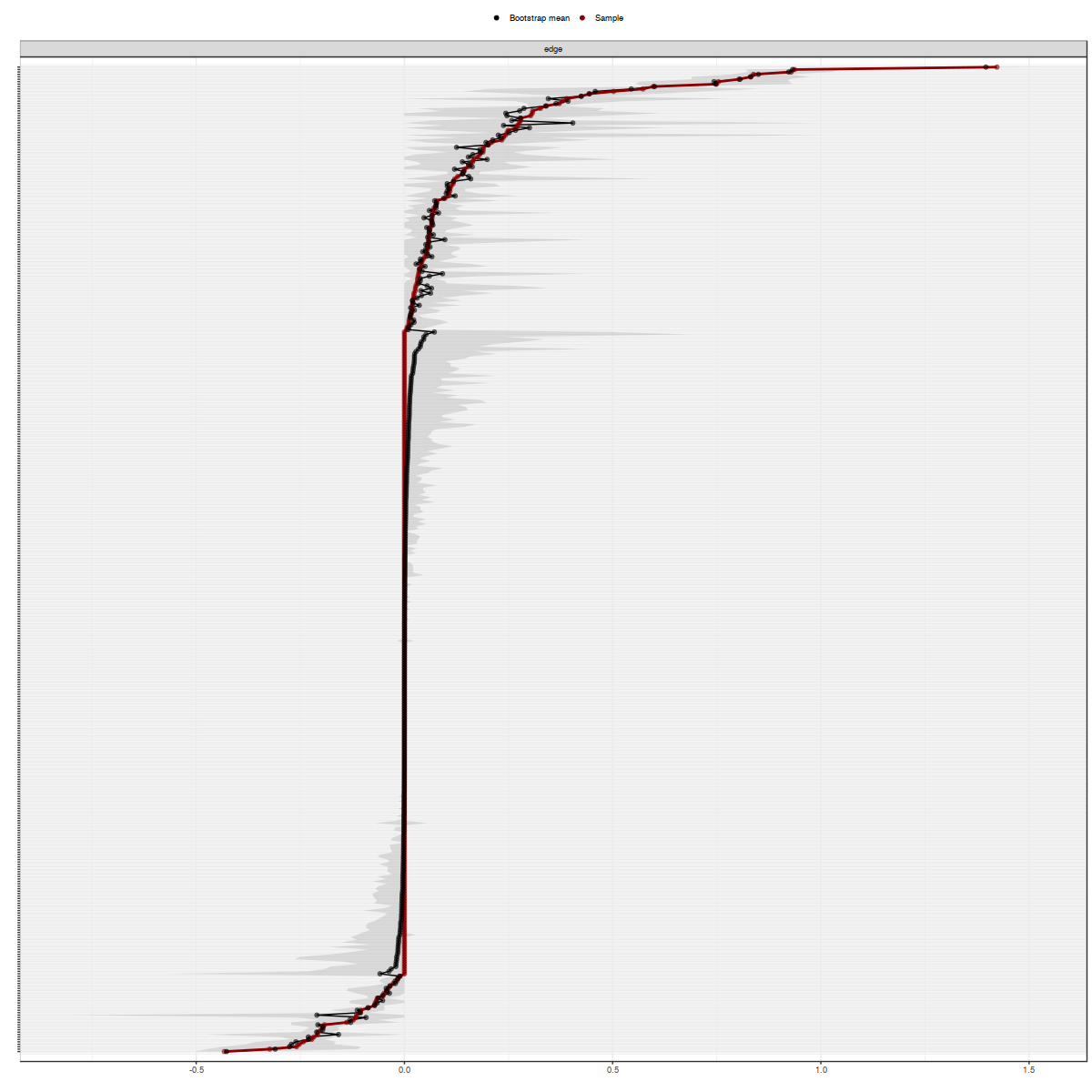

Figure S1: bootstrapped confidence intervals of the edge weights for model 1. The x-axis indicates the edge weights and the y-axis indicates the edge.

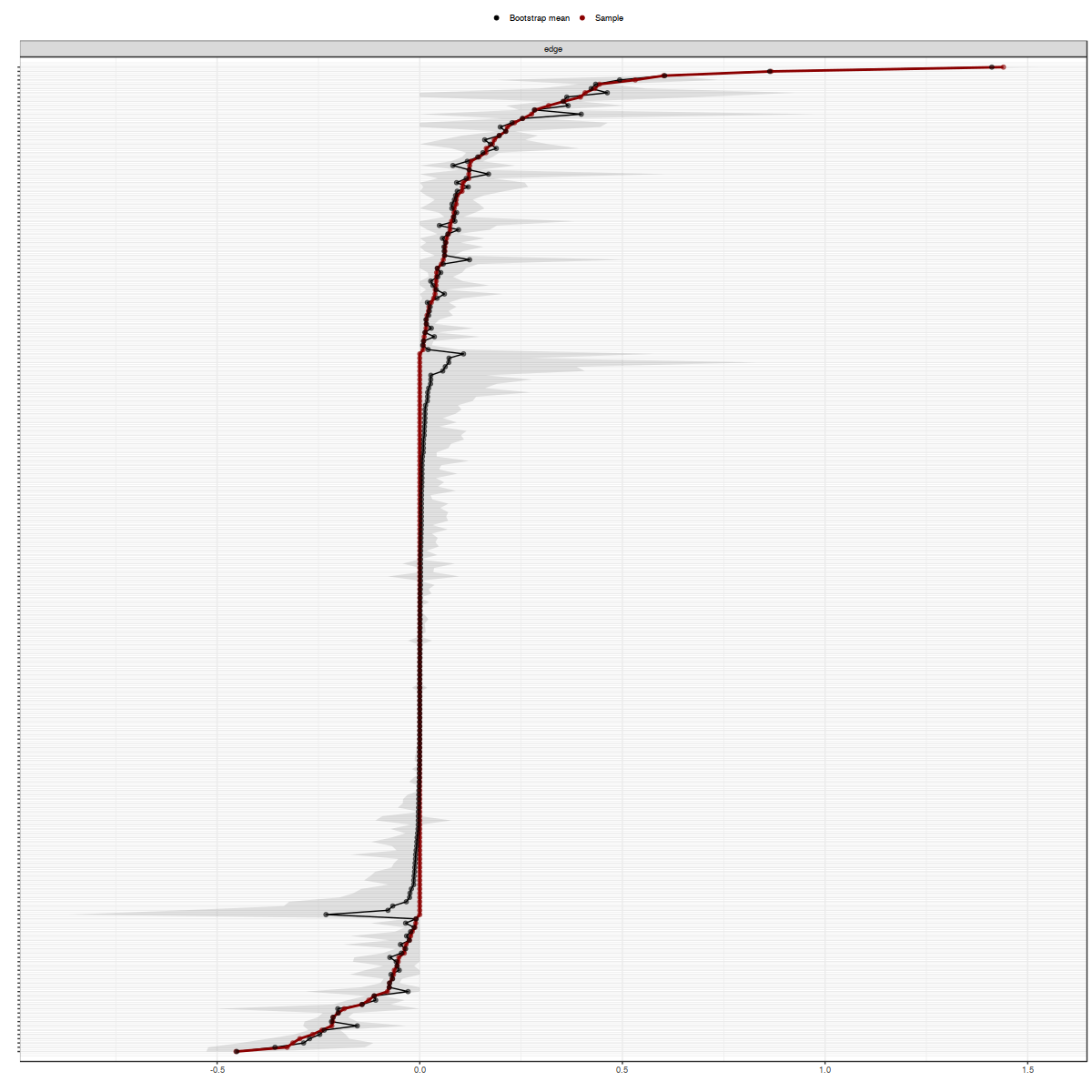

Figure S2 bootstrapped confidence intervals of the edge weights for model 2. The x-axis indicates the edge weights and the y-axis indicates the edge.

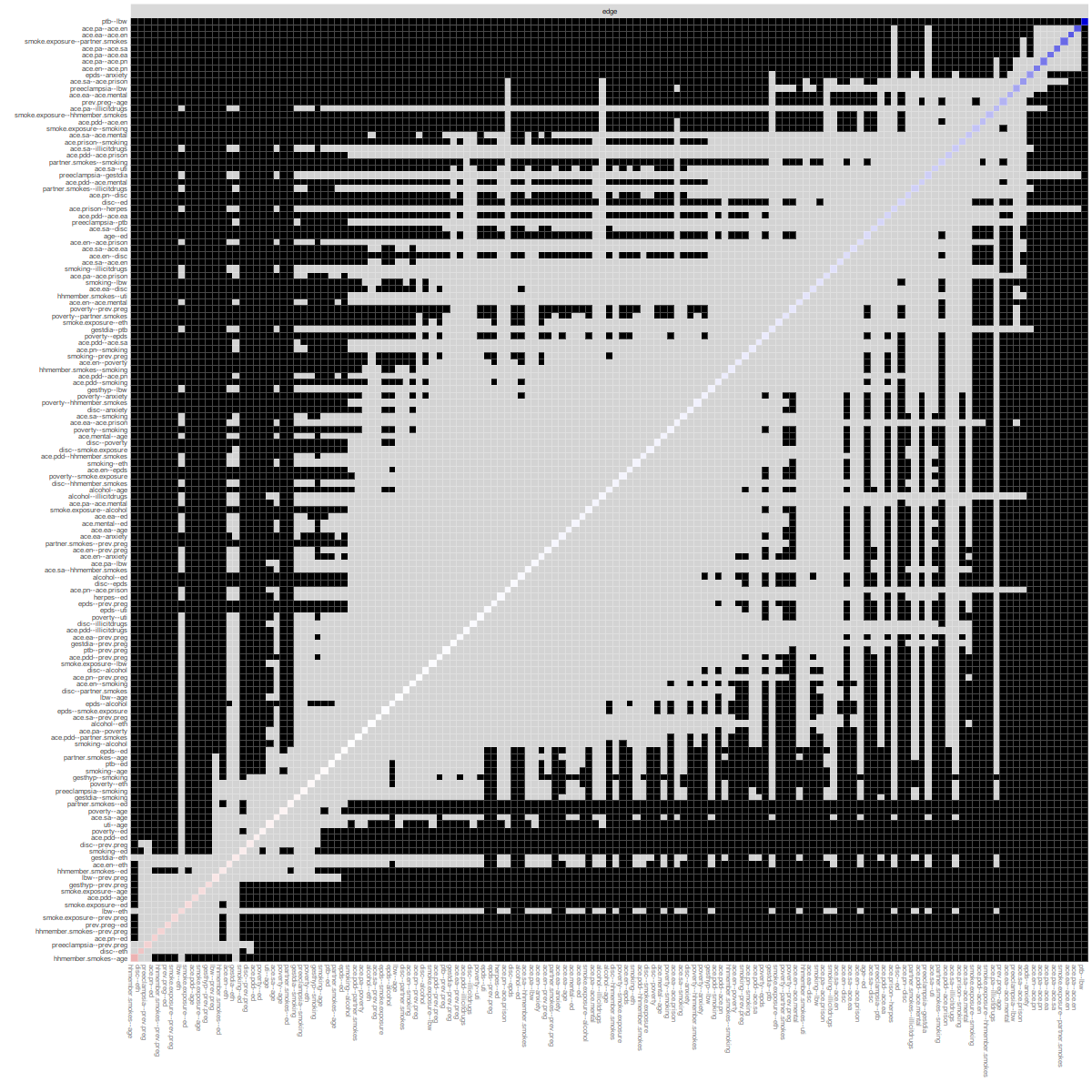

Figure S3: Results of the edge weight difference test for Model 1. Black squares indicate significant differences between two contrasted edge weights. Gray squares indicate difference between two edge weights which are non-significant.

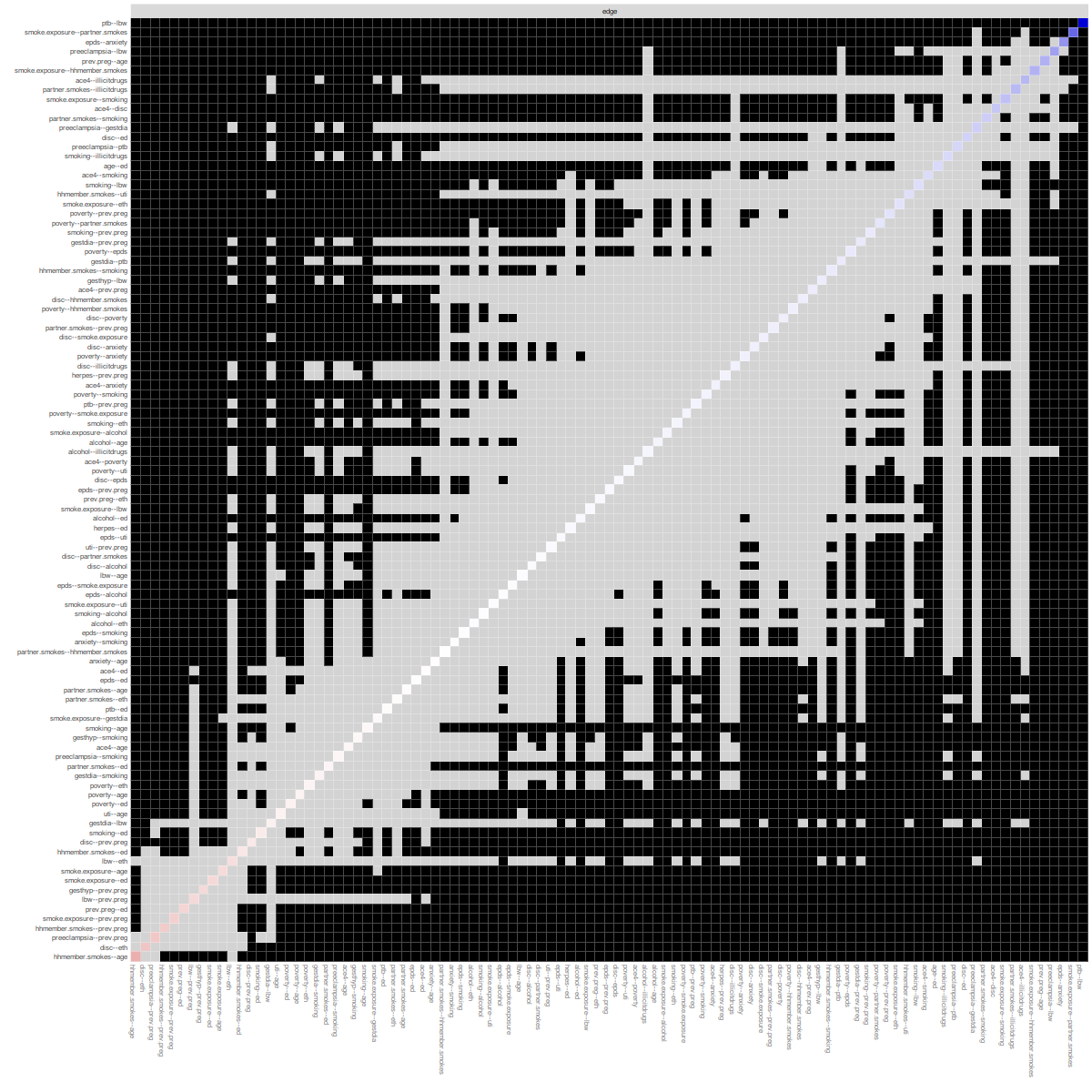

Figure S4: Results of the edge weight difference test for Model 2. Black squares indicate significant differences between two contrasted edge weights. Gray squares indicate difference between two edge weights which are non-significant.
