## Additional file 2 for "Using network analysis to illuminate the intergenerational transmission of adversity in the ALSPAC cohort"

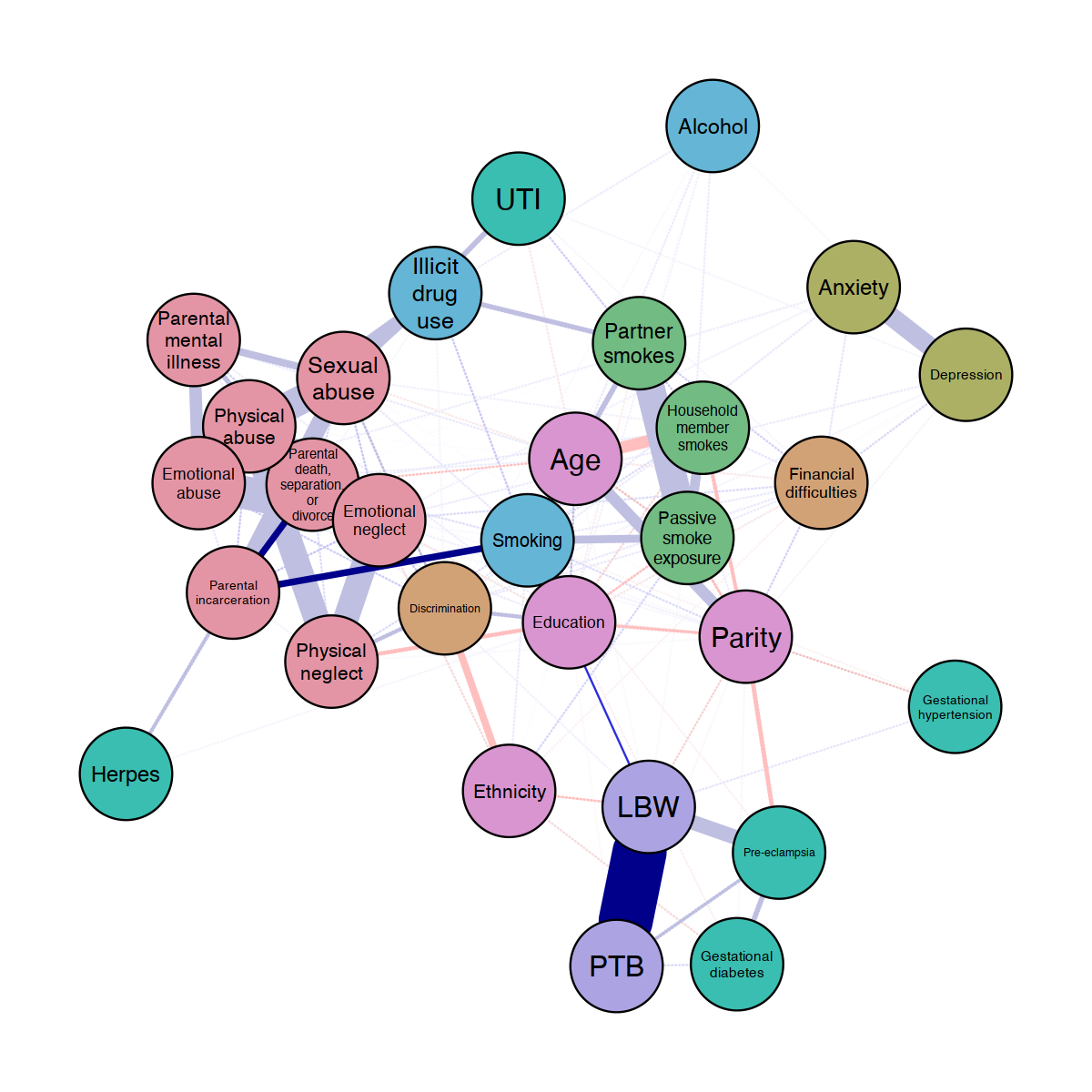

Figure S1: Network displaying the shortest pathway from Parental death, separation, or divorce node to infant low birthweight (LBW) and preterm birth (PTB) nodes. Blue edges suggest positive association while red edges represent negative associations.

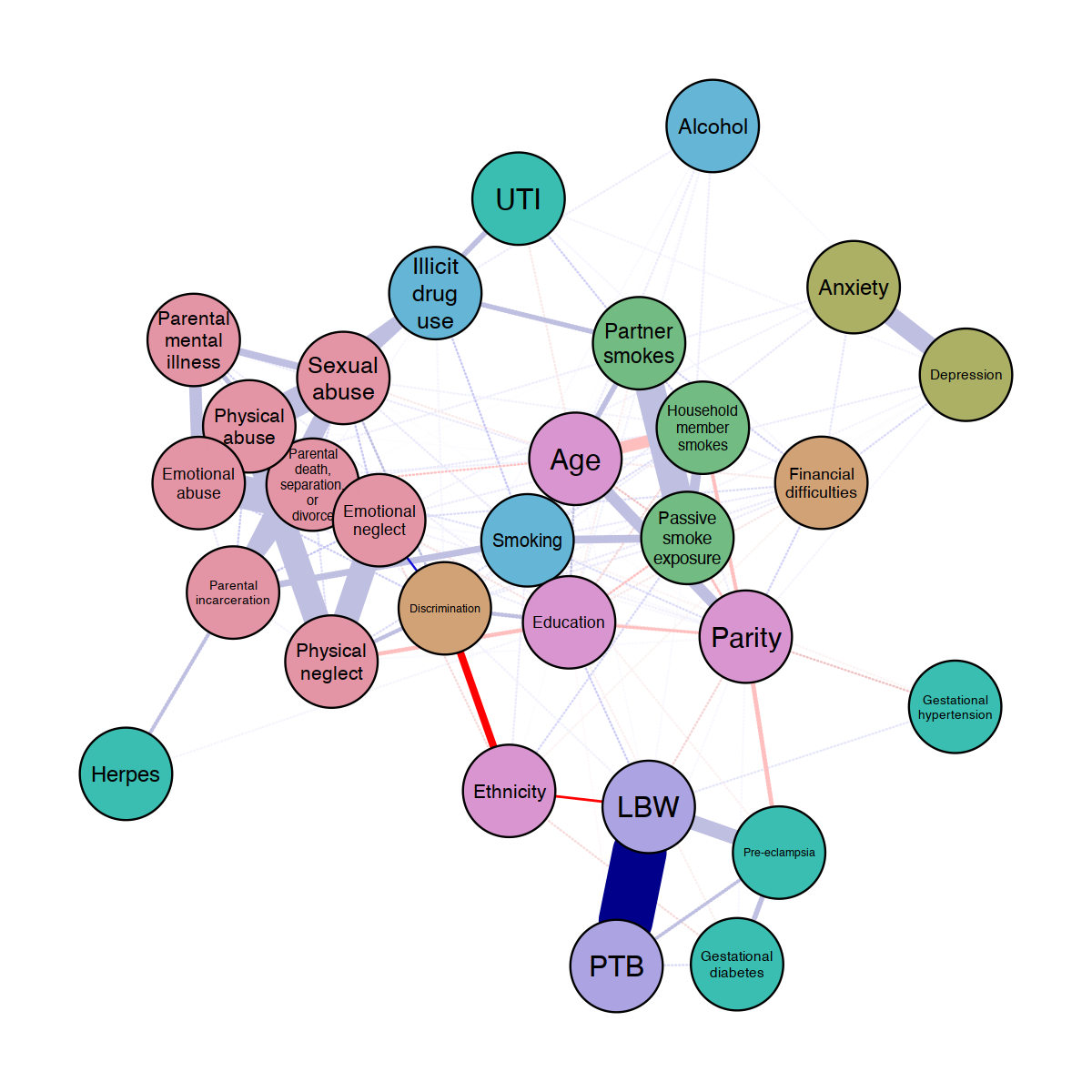

Figure S2: Network displaying the shortest pathway from Emotional neglect node to infant low birthweight (LBW) and preterm birth (PTB) nodes. Blue edges suggest positive association while red edges represent negative associations.

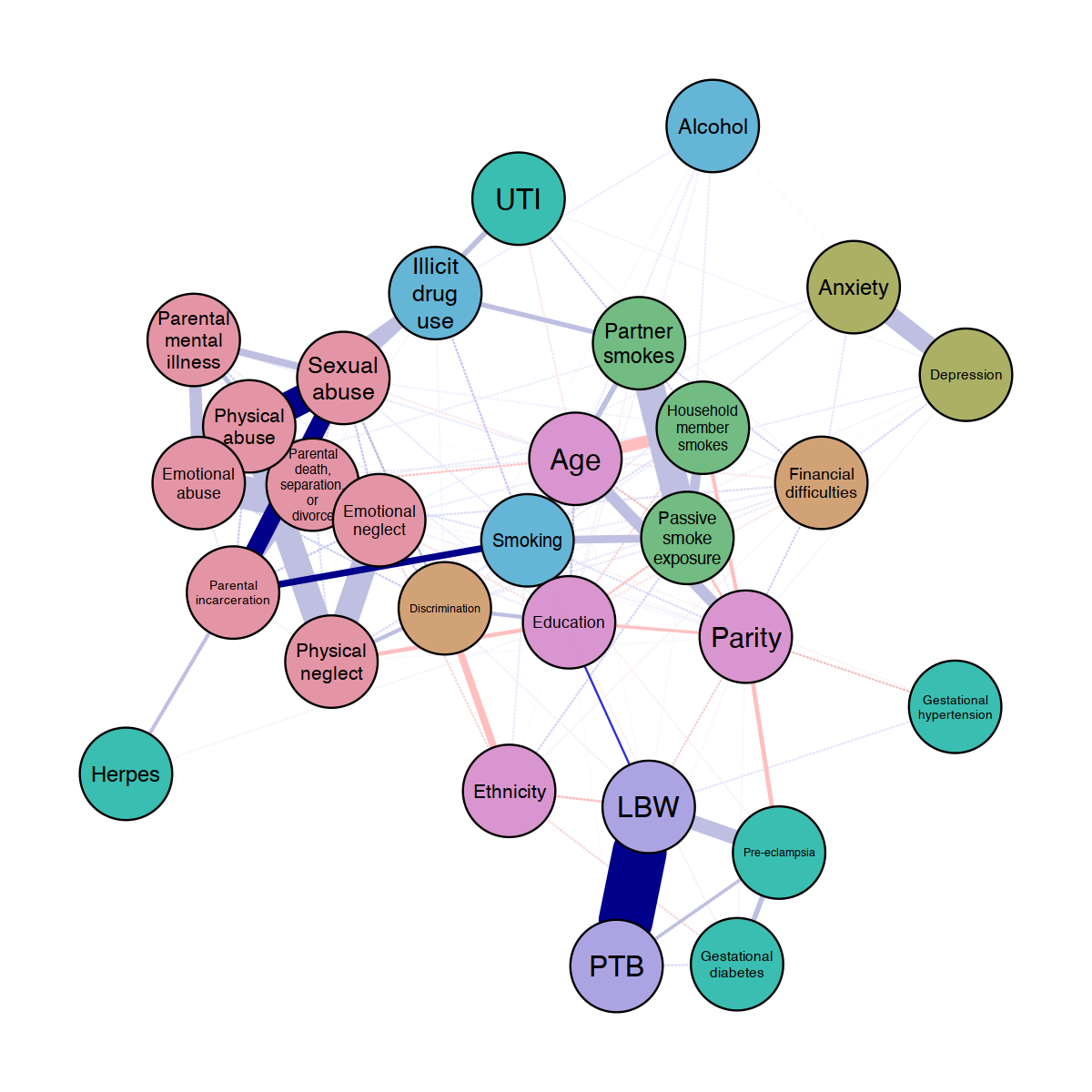

Figure S3: Network displaying the shortest pathway from Physical abuse node to infant low birthweight (LBW) and preterm birth (PTB) nodes. Blue edges suggest positive association while red edges represent negative associations.

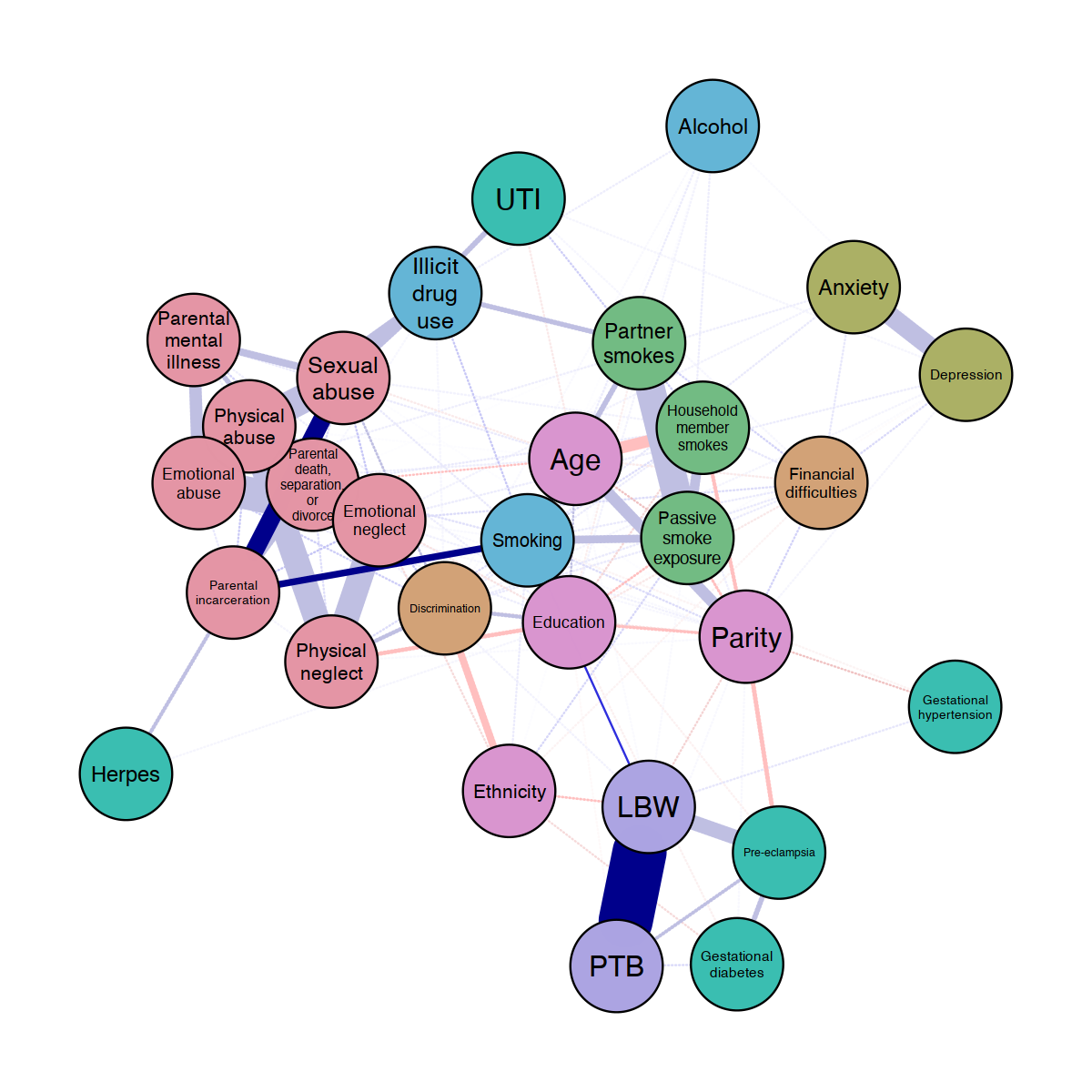

Figure S4: Network displaying the shortest pathway from Sexual abuse node to infant low birthweight (LBW) and preterm birth (PTB) nodes. Blue edges suggest positive association while red edges represent negative associations.

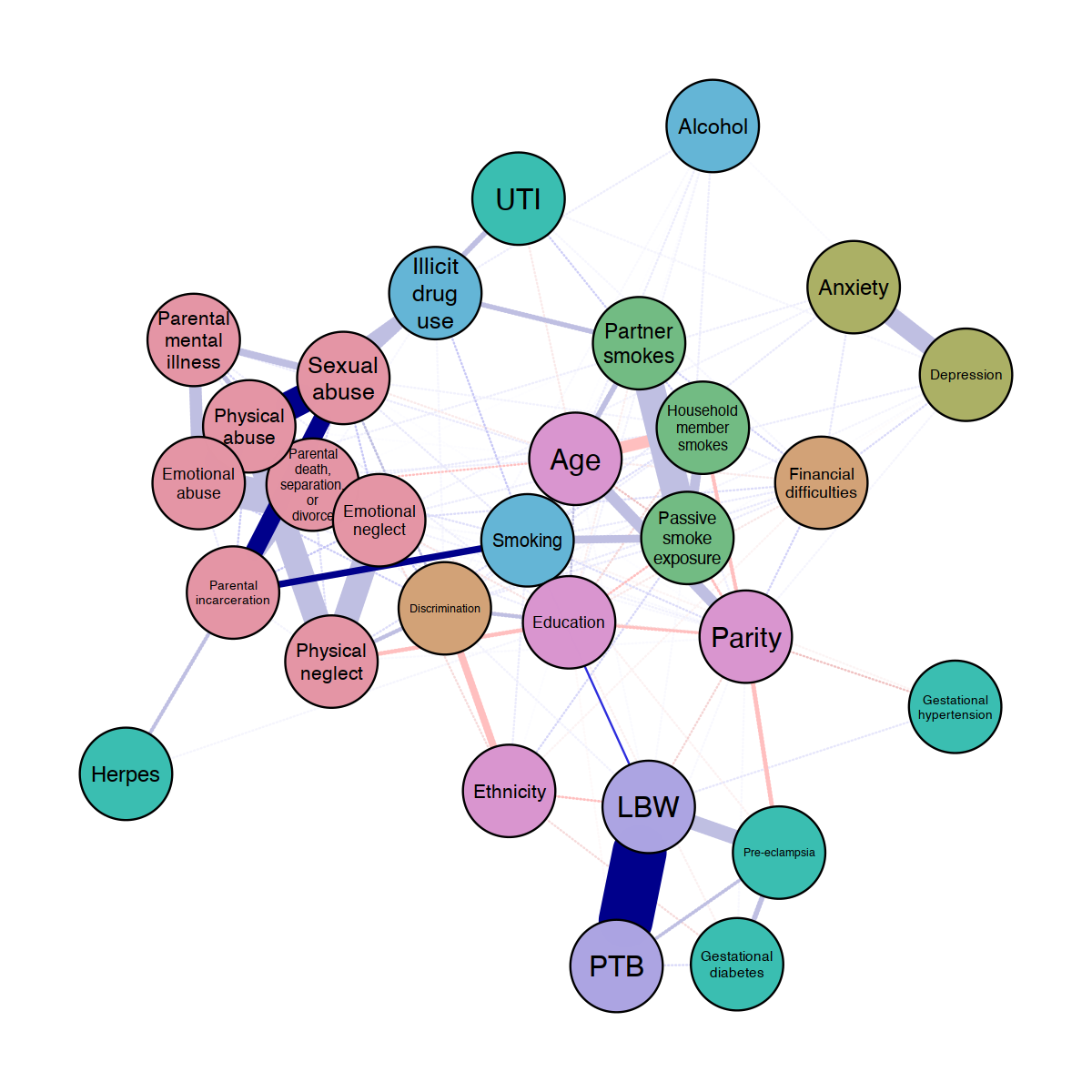

Figure S5: Network displaying the shortest pathway from Emotional abuse node to infant low birthweight (LBW) and preterm birth (PTB) nodes. Blue edges suggest positive association while red edges represent negative associations.

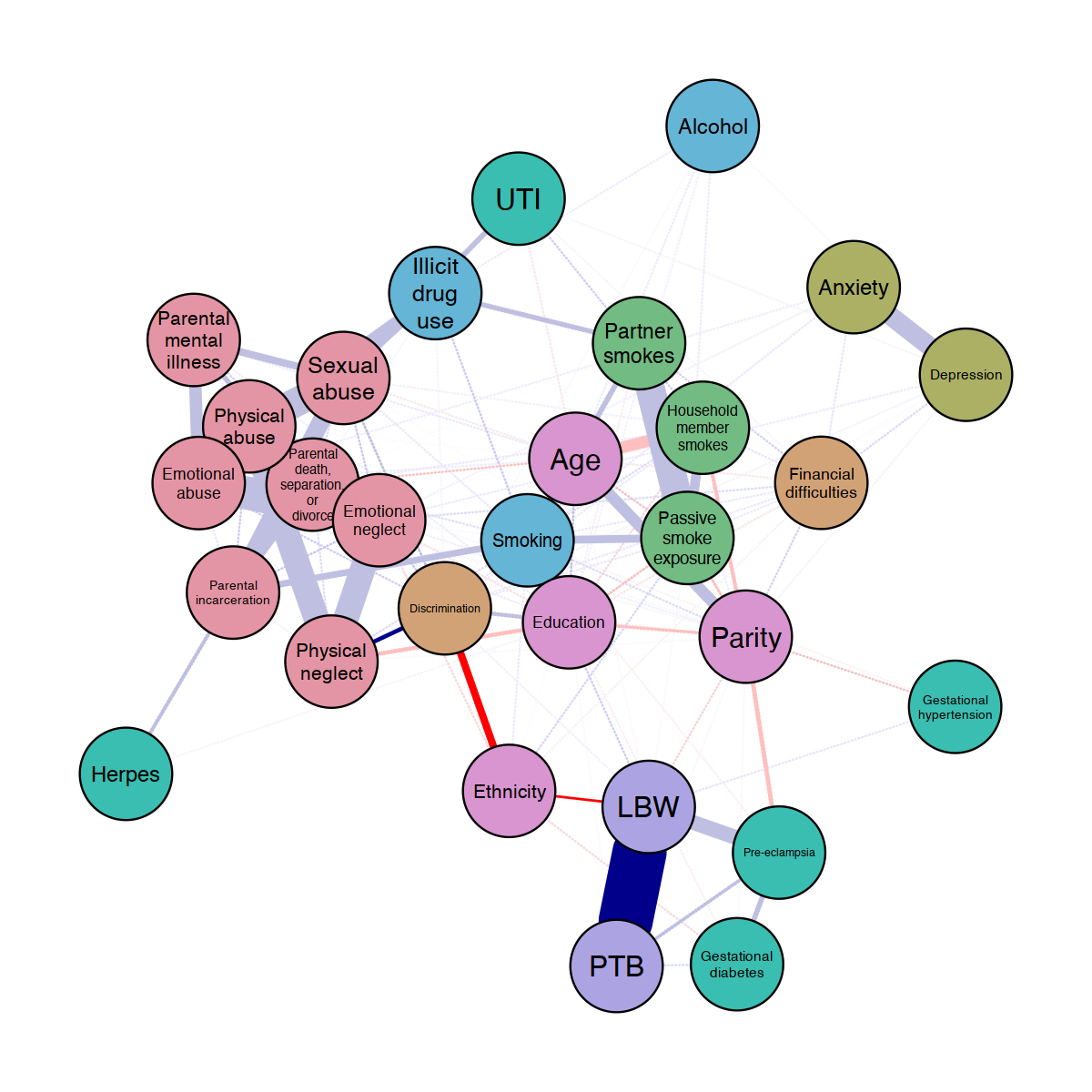

Figure S6: Network displaying the shortest pathway from Physical neglect node to infant low birthweight (LBW) and preterm birth (PTB) nodes. Blue edges suggest positive association while red edges represent negative associations.

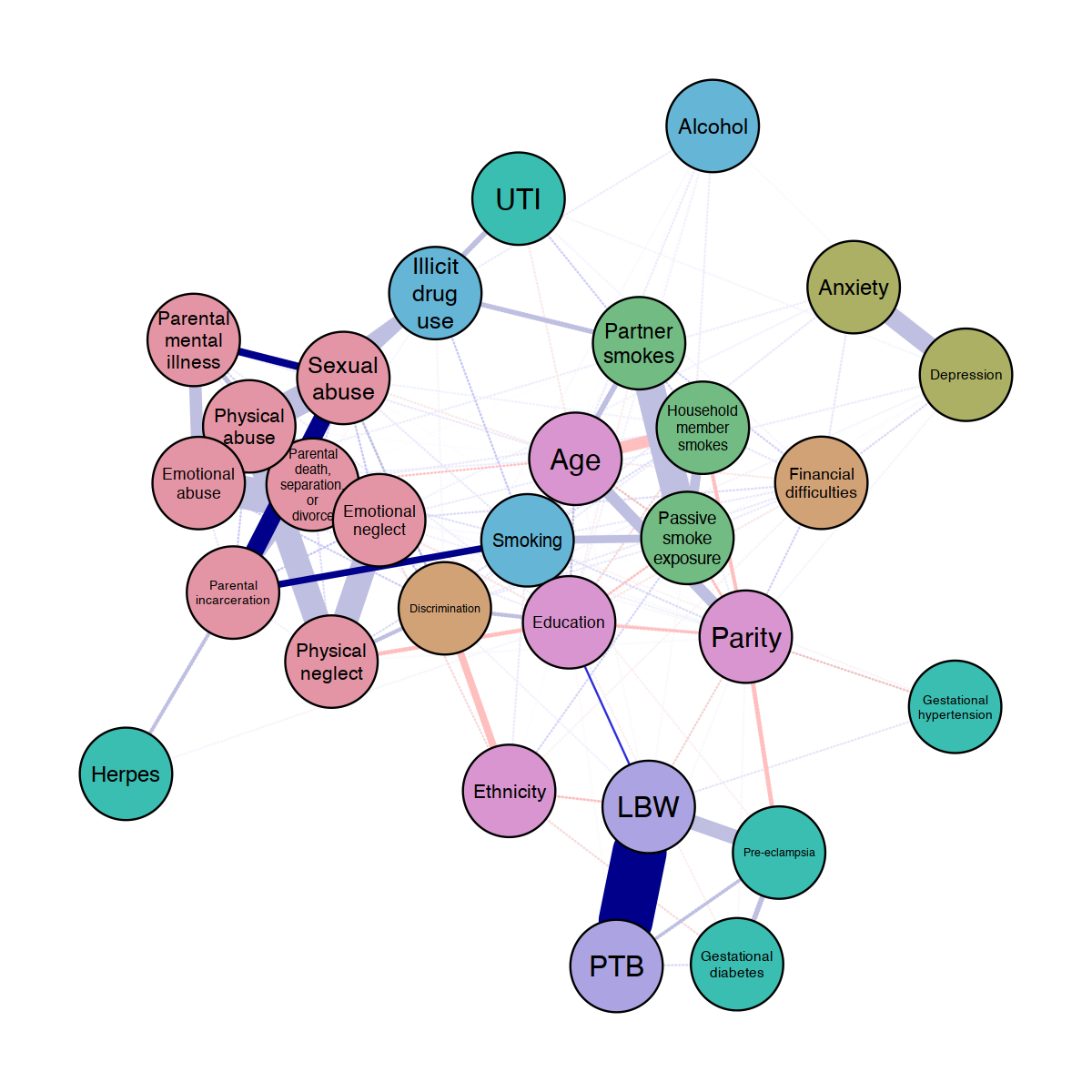

Figure S7: Network displaying the shortest pathway from Parental mental illness node to infant low birthweight (LBW) and preterm birth (PTB) nodes. Blue edges suggest positive association while red edges represent negative associations.

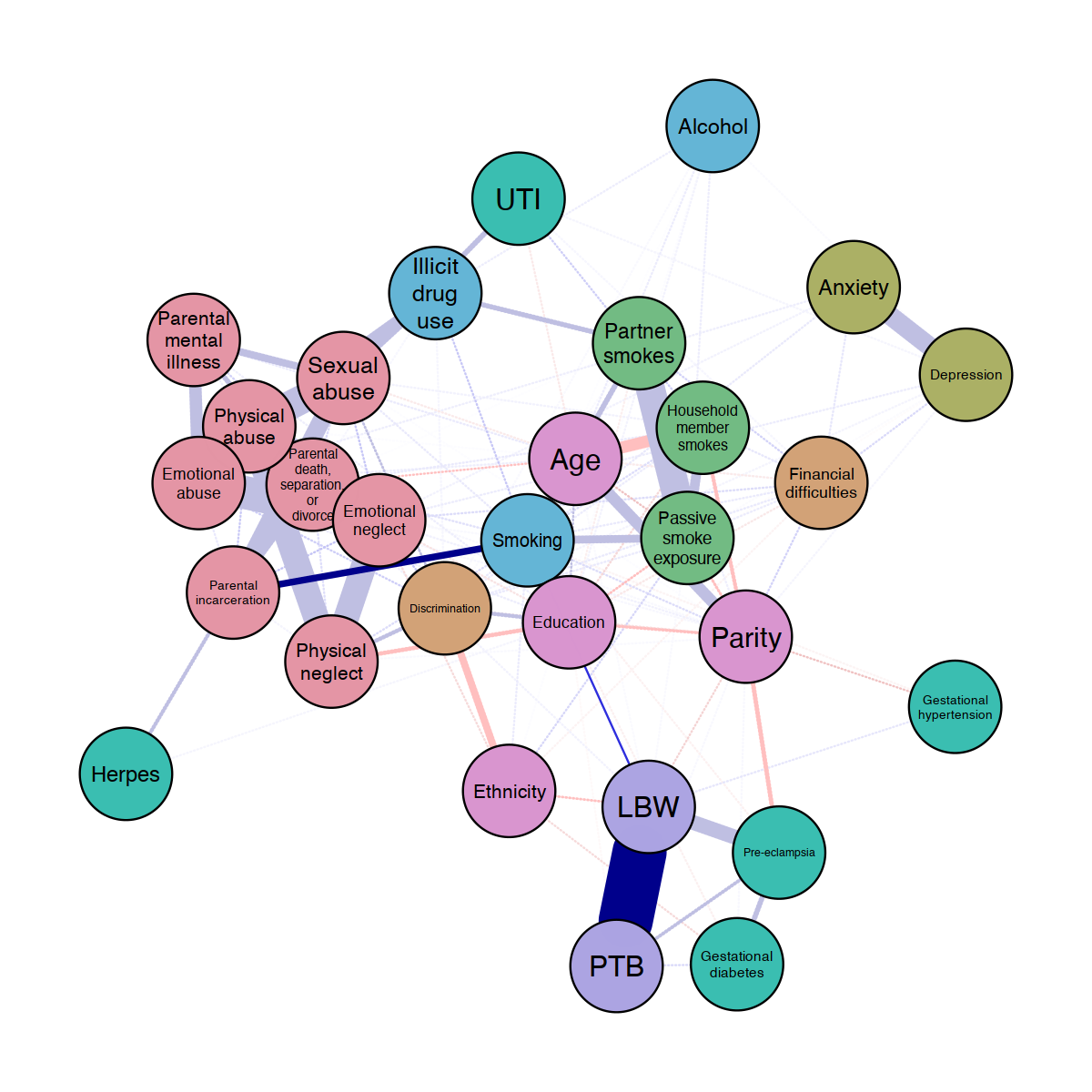

Figure S8: Network displaying the shortest pathway from Parental incarceration node to infant low birthweight (LBW) and preterm birth (PTB) nodes. Blue edges suggest positive association while red edges represent negative associations.

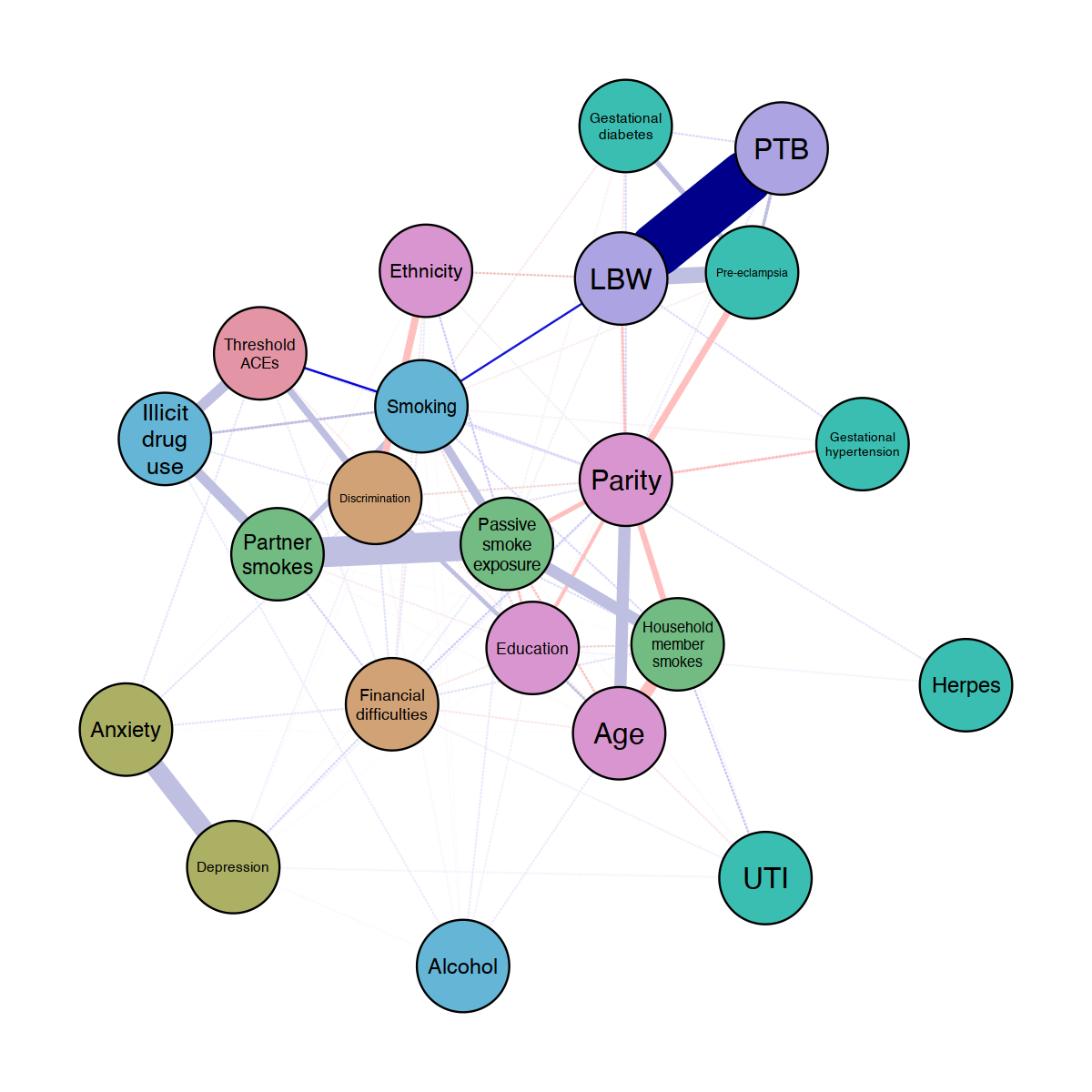

Figure S9: Network displaying the shortest pathway from threshold ACEs node to infant low birthweight (LBW) and preterm birth (PTB) nodes. Blue edges suggest positive association while red edges represent negative associations.

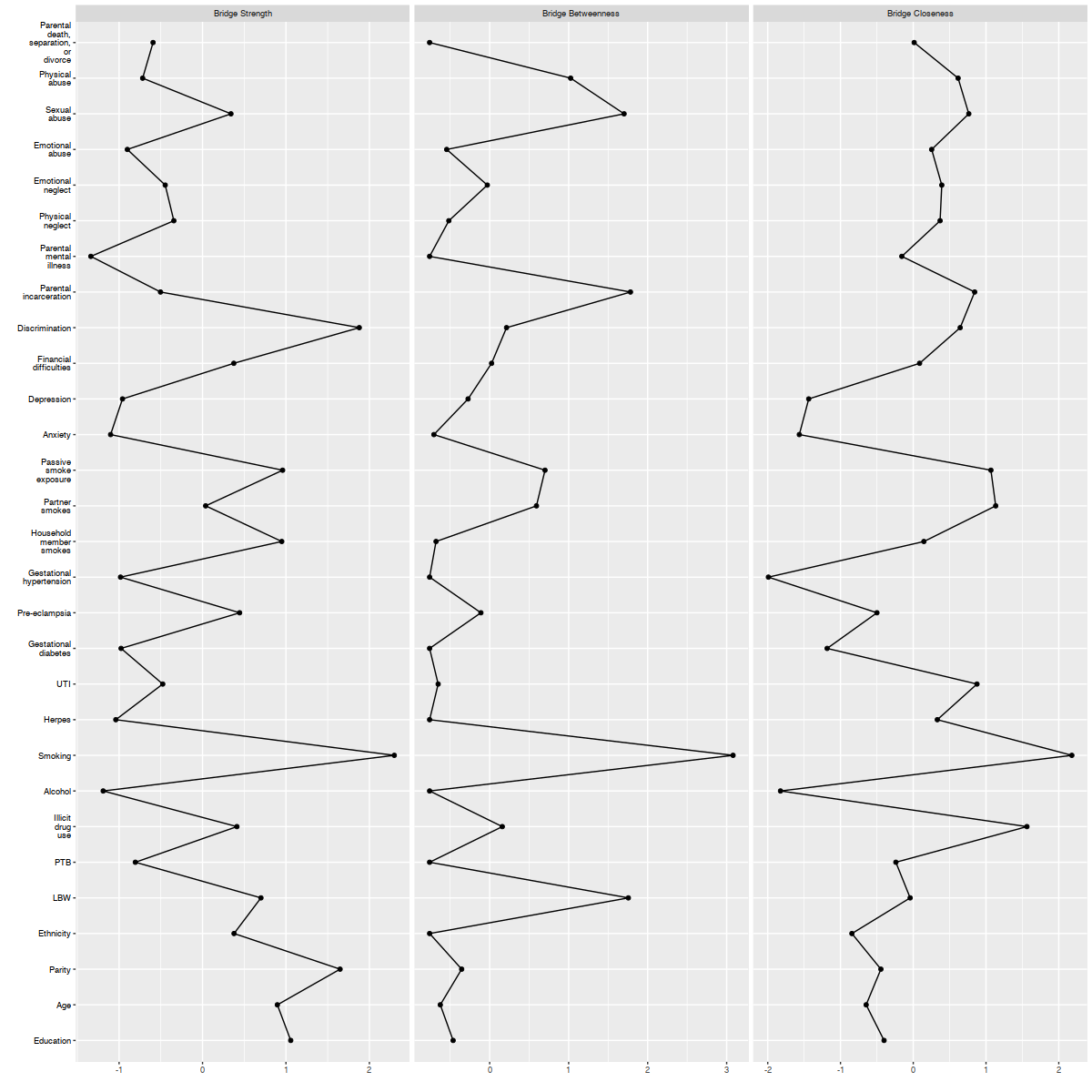

Figure S10: Z-transformed bridge centrality indices of model 1. Column 1 plots bridge strength, column 2 plots bridge betweenness, and column 3 plots bridge closeness.

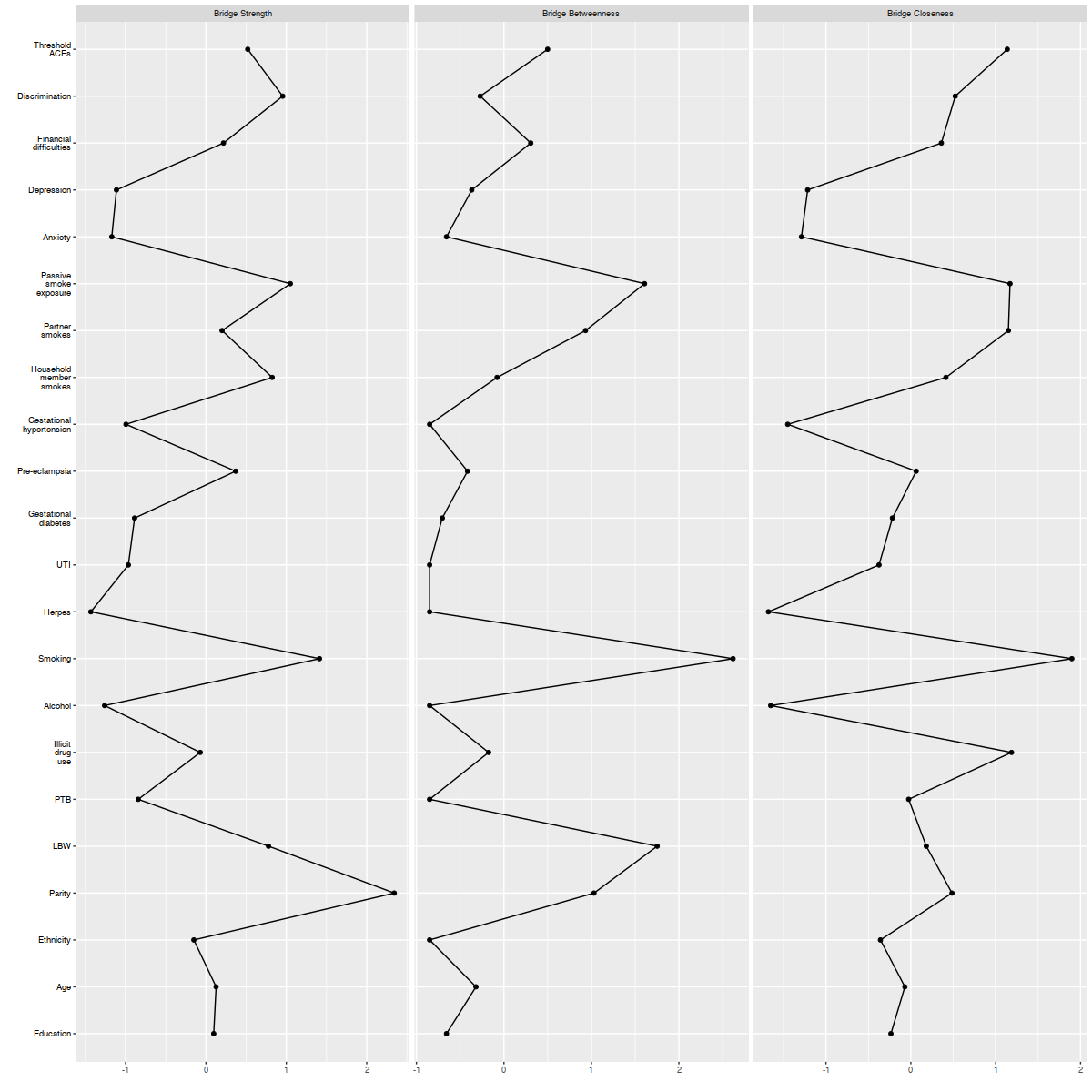

Figure S11: Z-transformed bridge centrality indices of model 2. Column 1 plots bridge strength, column 2 plots bridge betweenness, and column 3 plots bridge closeness.
